# A baseline transcriptional signature associates with clinical malaria risk in RTS,S/AS01-vaccinated African children

**DOI:** 10.1101/2021.05.19.21257227

**Authors:** Gemma Moncunill, Jason Carnes, William Chad Young, Lindsay N. Carpp, Stephen De Rosa, Joseph J. Campo, Augusto J. Nhabomba, Maximillian Mpina, Chenjerai Jairoce, Greg Finak, Paige Haas, Carl Murie, Phu Van, Héctor Sanz, Sheetij Dutta, Benjamin Mordmüller, Selidji T. Agnandji, Núria Díez-Padrisa, Nana A. Williams, John J. Aponte, Clarissa Valim, Daniel E. Neafsey, Claudia Daubenberger, Juliana McElrath, Carlota Dobaño, Ken Stuart, Raphael Gottardo

## Abstract

In a phase 3 trial in African infants/children, the RTS,S/AS01 (GSK) vaccine showed moderate efficacy against clinical malaria. We aimed to identify RTS,S/AS01-induced signatures associated with clinical malaria by analyzing antigen-stimulated peripheral blood mononuclear cells sampled from a subset of trial participants at baseline and month 3 (one month post-third dose). RTS,S/AS01 vaccination was associated with downregulation of B-cell and monocyte-related blood transcriptional modules (BTMs) and upregulation of T-cell related BTMs, as well as higher month 3 (vs baseline) circumsporozoite protein (CSP)-specific CD4^+^ T-cell responses. There were few RTS,S/AS01-associated BTMs whose month 3 levels correlated with malaria risk. In contrast, baseline levels of BTMs associated with dendritic cells and with monocytes (among others) correlated with malaria risk. A cross-study analysis supported generalizability of the baseline dendritic cell- and monocyte-related BTM correlations with malaria risk to healthy, malaria-naïve adults, suggesting inflammatory monocytes may inhibit protective RTS,S/AS01-induced responses.

## Introduction

Malaria remains a serious public health problem, with an estimated 229 million cases and 409,000 related deaths in 2019 (1). Despite the strides that interventions such as long-lasting insecticide treated bed nets, improved vector control and diagnostic tests, and mass antimalarial drug administration campaigns have made towards reducing malaria-related morbidity and mortality (2, 3), there is a critical need for an effective malaria vaccine (4, 5).

The RTS,S/AS01 malaria vaccine targets the pre-erythrocytic stage of the parasite life cycle and has been designed to elicit strong humoral and cellular immune responses against the *Plasmodium falciparum* circumsporozoite protein (CSP) (6). This recombinant vaccine consists of a protein containing multiple immunodominant NANP repeats and the carboxy terminus of CSP fused to hepatitis B virus surface antigen (HBs) formulated in the AS01 adjuvant (7).

In a phase 3 trial in 15,459 African infants and children (ClinicalTrials.gov NCT00866619) (8–11), RTS,S/AS01 demonstrated 56% vaccine efficacy (VE) against clinical malaria (follow up time: 12 months post-last dose) in children aged 5-17 months at enrollment and 31% in infants aged 6-12 weeks at enrollment. In 2015, RTS,S/A01 became the first malaria vaccine to receive a positive opinion by the European Medicines Agency under Article 58 (12), and it was recommended by the World Health Organization for a malaria vaccine pilot implementation program in Ghana, Malawi, and Kenya that started in 2019 (13).

A critical limitation of the RTS,S vaccine is that VE is moderate (lower in infants than children) and wanes substantially within the first 18 months (10). The identification of immune correlates of protection could help guide iterative vaccine improvements and expedite vaccine evaluation. Excellent work has been done on elucidating correlates of RTS,S/AS01-mediated protection in healthy, malaria-naïve adults using the controlled human malaria infection (CHMI) model (14–20), and cohort studies in African infants and children have implicated vaccine-induced anti-CSP antibodies (21–23), as well as CSP-specific Th1 cytokines (24), and CD4^+^ T cells (albeit to a lesser extent) in protection in this population (reviewed in (25)).

The MAL067 study, an ancillary study to the RTS,S/AS01 phase 3 trial, was conducted to address key knowledge gaps of RTS,S induced immune responses and their correlation with protection against field challenge. Using RNA-sequencing (RNA-seq) data from antigen- or vehicle-stimulated PBMC obtained at baseline and one month post-final dose from infants and children enrolled in Bagamoyo, Tanzania and Manhiça, Mozambique, we aimed to identify baseline and/or RTS,S/AS01-induced signatures associated with clinical malaria risk. Post-vaccination anti-CSP antibody titers, cytokine profiles, and T-cell responses, the latter of which were additionally assessed in samples from participants enrolled in Lambaréné, Gabon, were also examined as correlates of clinical malaria and/or of RTS,S/AS01-induced transcriptional responses.

The major finding of our study is that pre-vaccination expression of immune-related blood transcriptional modules (BTMs), including BTMs related to dendritic cells and monocytes, correlated positively with malaria risk in RTS,S/AS01-vaccinated African children; moreover, this dendritic cell- and monocyte-related signature appeared to be generalizable to malaria-naïve RTS,S/AS01-vaccinated healthy adults.

## Materials and Methods

### MAL067 trial

During the MAL055 Phase 3 study (ClinicalTrials.gov identifier NCT00866619; (11)), infants (6-12 weeks) and children (5-17 months) received RTS,S/AS01 or comparator (rabies vaccine for children; meningococcal C conjugate vaccine for infants), with injections given at month 0 (baseline), month 1, and month 2 (Figure 1). MAL067 was a multi-center immunology ancillary study nested within MAL055 and selection of participants for MAL067 is described in (24). Of the seven trial sites included in MAL067, PBMC samples were collected at baseline (only children) and again at month 3 (one-month post-third dose) at three. The present study analyzes PBMC data from children in Bagamoyo (Tanzania) and from infants and children in Manhiça (Mozambique), as well as from infants and children in Lambaréné (Gabon) (only intracellular cytokine staining (ICS)/immunophenotyping for the latter). All malaria cases with samples collected were included together with 2 to 3 matched non-malaria controls for each malaria case, prioritizing subjects who had samples at both month 0 and month 3 and in whom the complete set of antigen stimulations was conducted.

**Figure 1:**
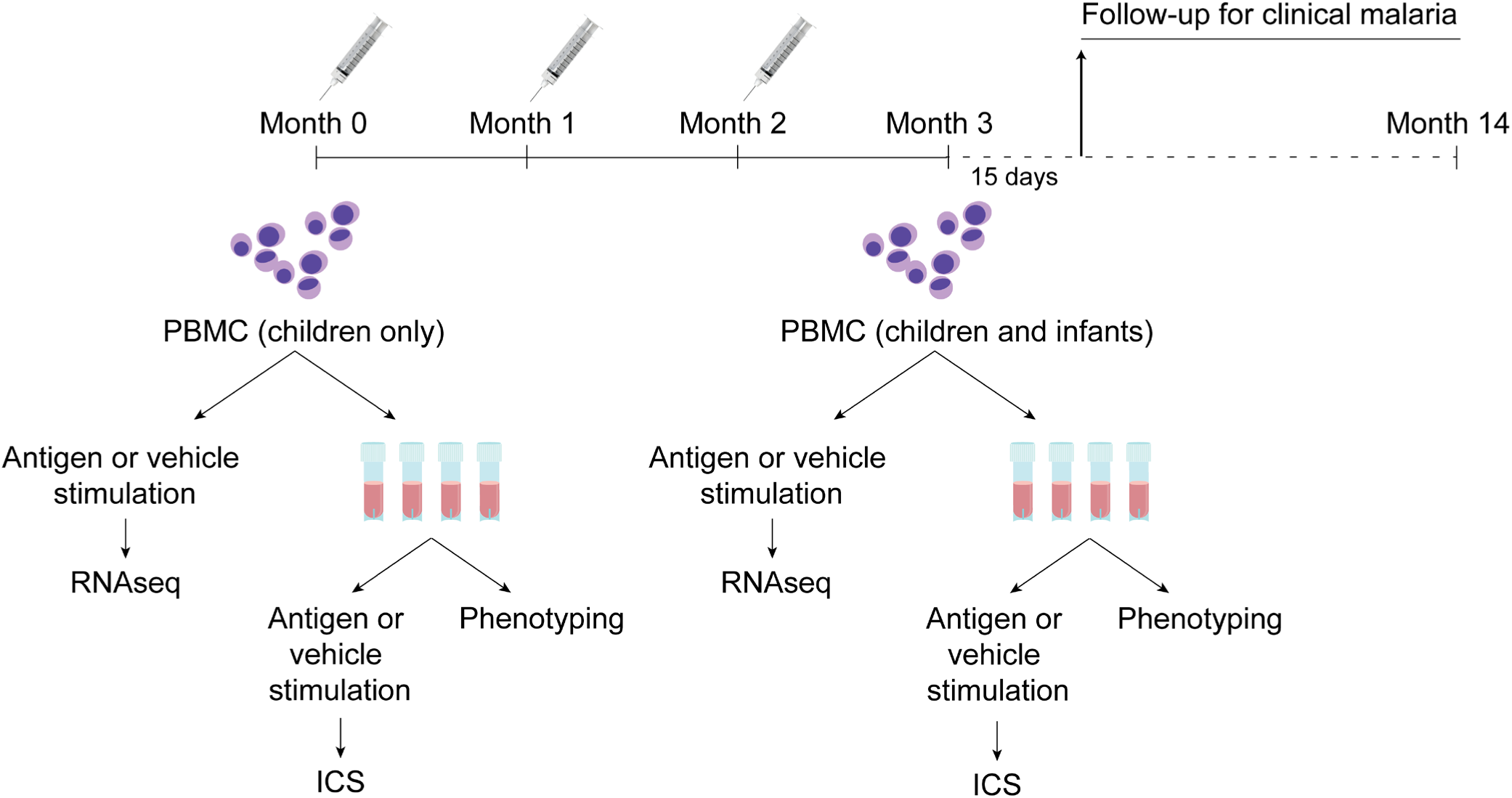
Schematic showing vaccination and sampling schedule. Participants received RTS,S/AS01 (or comparator) at month 0, month 1, month 2; peripheral blood mononuclear cells (PBMC) were collected for RNA-sequencing at month 0 and month 3 (1 month post-final dose). Stim, stimulation.

### Case-control definitions

Cases were defined as participants who had any episode of clinical malaria (fever >37.5°C with any parasitemia by blood smear) in the 12 months of follow up after month 3.5, identified by passive case detection (subjects who sought care at a health facility). Controls were participants who did not have any clinical malaria case during the 12 months of follow-up. Controls were matched to cases based on site, age group, and time of vaccination and follow-up. Up to 3 controls were selected for each case among RTS,S/AS01 vaccinees, and 2 controls were selected for each case among comparator vaccinees.

### PBMC collection and antigen stimulation

PBMC collection and stimulation with DMSO (vehicle control), apical membrane antigen (recombinant AMA1, FVO strain), CSP (peptide pools), or HBS (peptide pools) is described in (24)). For stimulation before RNA extraction, 4×10^5^ freshly isolated PBMC seeded in duplicates were rested for 12 h and then incubated 12 h at 37°C with 1 μg/mL antigens in 96 well-plates. Plates were then centrifuged for 5 min at 250*g* at room temperature and cell pellet duplicates were resuspended and pooled in RLTbuffer at Bagamoyo or RNAprotect (Qiagen) at Manhiça, transferred into a 96 well V-bottomed plate (Kisker, AttendBio G096-VB) and sealed with adhesive foil (Kisker, Attend Bio G071-P) and cryopreserved at −80°C till RNA extraction.

For stimulation before ICS, cryopreserved PBMC were thawed and then rested in a 37°C, 5% CO_2_ incubator overnight. PBMC were stimulated for 6 h with the same peptide pools as above, DMSO (vehicle control), and Staphylococcal enterotoxin B as a positive control.

### Flow immunophenotyping

Leftover cryopreserved PBMC thawed for ICS (0.5-1×10^6^ cells) were used for leukocyte phenotyping. The flow cytometry panel and staining protocol used is described in (26). Data were acquired using a BD LSR II flow cytometer (BD Biosciences) directly from 96-well plates using a high throughput sampler. Flow cytometry analysis was performed using FlowJo software (Version 9.9 Tree Star). The gating strategy was performed as in (26, 27).

### Intracellular cytokine staining

Antigen-or vehicle-stimulated PBMC were stained using a flow cytometry panel and protocol previously described (27, 28) with the additional marker IL-13. Antibody details can be found in Table S1. Data was acquired and analyzed as above.

### RNA isolation and sequencing

RNA was extracted at the Center for Infectious Disease Research (CIDR, Seattle) using the Promega SV 96 RNA Isolation kit following the manufacturer’s protocol. Samples kept in RNAprotect were centrifuged at 4,000g for 7 minutes at 4°C, cell pellets were resuspended in 150 μL RLT Buffer, and 150 μL of 70% ethanol was added prior to processing with Promega SV 96 RNA Isolation kit. RNAs were eluted with 100 μL nuclease free water. Each 96-well extraction batch was spot checked by Bioanalyzer using Agilent RNA 6000 Pico chip and had an average RIN score of 7.4. RNA samples were distributed in 384 well plates for library preparation. Samples from the same individuals were in the same plate and key study variables (vaccine, site and cases-controls) were checked for balance across plates to avoid batch effects.

An optimized version of Digital Gene Expression (DGE) was used, further reducing the reverse transcriptase reaction volume. In brief, poly(A)+ mRNA from antigen-stimulated PBMCs was linked to unique molecular identifiers (UMIs) using a template-switching reverse transcriptase. Then, cDNA from multiple cells were pooled, amplified, and prepped for multiplexed sequencing using a transposon-based fragmentation method, enriching for 3′ ends and preserving strand information. Nextera XT library preparation and sequence paired-ends were employed on an Illumina NextSeq instrument at the Broad Institute.

### Immunogenicity data analyzed for correlations with BTM expression

#### Antibody

NANP-specific, HBS-specific, and C terminal domain of CSP (C-term)-specific antibody data from previous studies were analyzed for correlations with BTM expression as described below. IgG titers (EU/mL) against NANP and against HBS were obtained from the MAL055 trial database (8–11). IgG titers (EU/mL) against NANP and C-term were measured by ELISA at IAVI-HIL (21). IgG and IgM levels (Median Fluorescence Intensity, MFI) against NANP, C-terminal CSP and HBS together with 35 RTS,S/AS01 vaccine-unrelated malaria antigens were measured by Luminex technology (22, 23).

#### Cellular

Data from cytokines in the supernatants of the same CSP and HBS PBMC stimulations analyzed by Luminex in a previous study (24) were used here for detecting BTMs correlated with RTS,S/AS01 immunogenic responses.

### Data processing and statistical analysis

#### Preprocessing

Preprocessing of RNAseq data was done by Broad Technology Labs. In brief, reads were aligned using BWA Aln version 0.7.10 using UCSC RefSeq (Human 19) with mitochondrial genes added. Quantified samples were then quality controlled using mapping summary statistics to remove low quality samples based on predetermined minimum values for the total number of mapped reads, percent of mapped reads mapped to the human genome, etc. Downstream analysis was applied only to reads that mapped uniquely to a UMI and only mapped to isoforms of the same gene (UMI.unq).

#### Normalization

The TMM normalization method (29) was applied to account for differing number of read counts and to address unwanted technical variation. The voom transformation (30) from the limma R package (31) was applied to standardize and appropriately weight the data for use in linear models.

#### Quality control

In a pilot study, we found that sample libraries that exhibit less than 75,000 total RNAseq reads per sample were of low quality. Thus, such libraries were removed from the study. Genes that had less than 20 samples (around 10%) with read counts greater than 5 were also removed. For further details, see the Supplementary Methods.

#### Differential expression

Differential expression was assessed using module-based (using voom and camera (32)) approaches as implemented in the limma package. Camera, combined with voom, is one of the few gene set enrichment analysis methods that can properly account for inter-gene correlation in RNA-seq data. Specifically, Camera estimates the variance inflation factor for the gene expression that results from inter-gene correlation in the data and incorporates it into test procedures to control the apparent false discovery rate. This step is important since significant correlation is expected among genes in the same module. Inference was based on p-values adjusted for multiple testing by controlling the false discovery rate with the Benjamini-Hochberg (33) method. Differential expression was used to down-select modules constituting the PBMC transcriptional response to RTS,S/AS01 vaccination comparing RTS,S/AS01 vaccinees with comparator at month 3 and pre- and post-RTS,S/AS01E vaccination in children (month 3 vs month 0). Noninformative modules (“TBA”) are omitted from all heatmaps.

Blood transcriptional module (BTM) analysis: BTMs used were from Li et al. (34). Resulting p-values across BTMs (within stimulation condition) were adjusted for multiple testing with a false discovery rate (FDR) cutoff of 0.2. Only these significant BTMs were tested as candidate immune correlates.

#### BTM correlations with immunogenicity

For each module, a score was calculated for each RTS,S recipient at month 3 and at month 0 based on the average normalized expression level of all genes in the modules, on the log scale. Spearman’s rank correlation was used to assess association between gene expression, functional antibody and cellular responses. Each correlation was tested (Pearson correlation test) and a p-value was obtained. P-values were adjusted within each response (across all gene sets); significance was defined as an adjusted p-value ≤0.2.

#### Correlates analysis

We identified BTMs significantly associated with protection using the limma package. The model adjusted for clinical and experimental covariates such as age and technical experimental factors. This model was applied to each BTM and stimulation condition identified in the down-selection process. Resulting p-values were adjusted for multiple testing with an FDR cutoff of 0.2. For the cross-study correlates analysis, BTMs were down-selected based on month 3 or month 0 (as appropriate) data from MAL067 (vehicle-stimulated PBMC only). In brief, month 3 or month 0 data for every BTM were tested and FDR adjustment was done across all BTMs. Only those BTMs with FDR < 0.2 in MAL067 were examined as potential correlates of challenge outcome in the CHMI studies, with FDR adjustment performed within each study.

#### Logistic regression

Conditional logistic regression was applied to assess which monocyte populations were predictive of clinical malaria. The clogit function from the R survival package was used and incorporated a stratified variable which grouped subjects according to a set of personal and clinical factors.

#### Analysis of antigen (Ag)-specific T cells

When analyzing Ag-specific T cells, vehicle-only stimulations (DMSO) were used to determine the effect of Ag stimulation over vehicle stimulation for each PBMC sample. The comparison was performed using the limma package (35) in R as follows: stimulation*vaccine, where stimulation = (HBS, AMA1, CSP) vs vehicle and vaccine = RTS,S/AS01 vs Comparator.

## Results

### Study population and sample collection scheme

PBMC RNA-seq data from a total of 193 participants were analyzed (Table 1). Baseline RNA-seq data were available for 36 recipients (8 cases, 28 controls) and 19 comparator recipients (5 cases, 14 controls), all of whom were children since baseline samples were not collected from infants. Month 3 RNA-seq data were available for 123 RTS,S/AS01 recipients (30 cases, 93 controls) and 70 comparator recipients (23 cases, 47 controls). All (100%) of the participants in Bagamoyo for whom Month 3 RNA-seq data were available were children, whereas nearly all (94.9%) in Manhiça were infants. Table S2 provides similar information for participants for whom immunophenotyping/ICS data were analyzed.

**Table 1:**
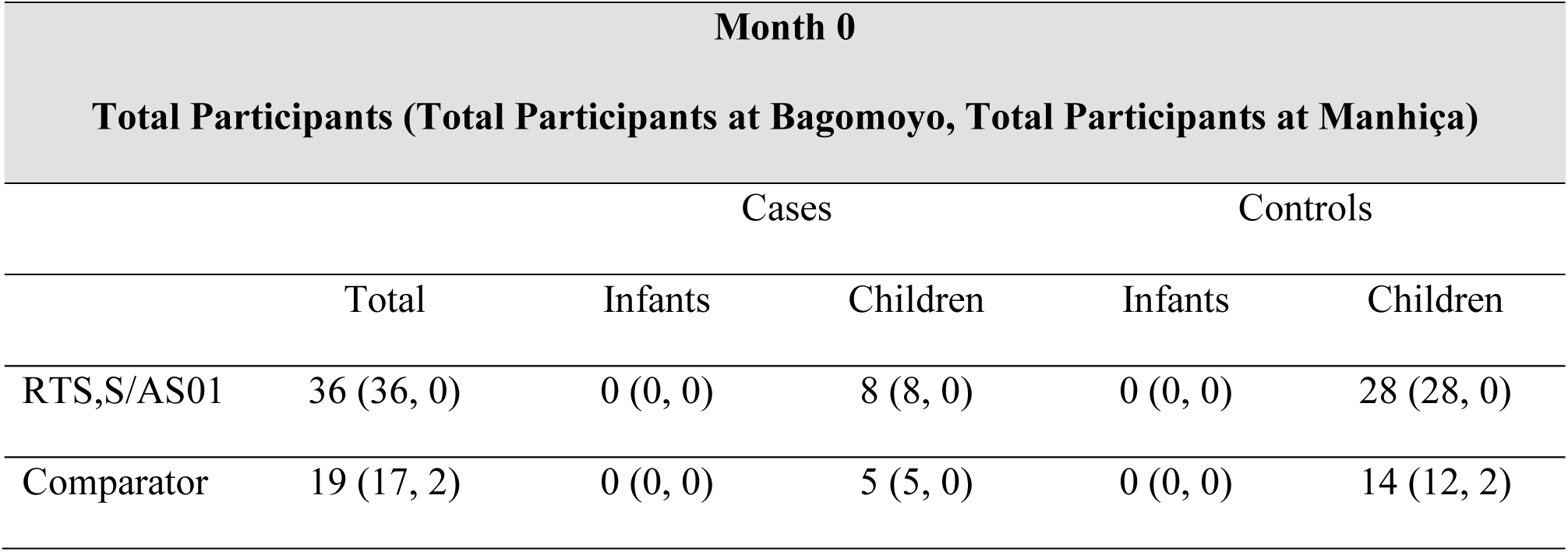
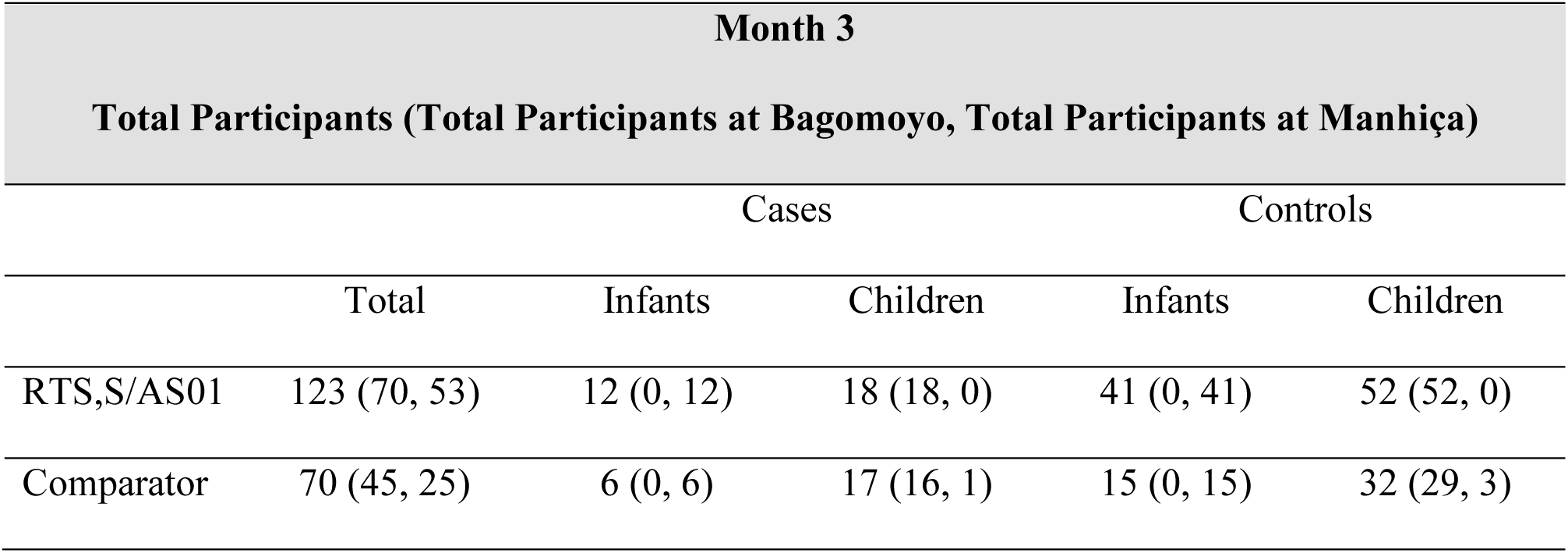
Numbers, age group, and case-control status of participants by site for whom PBMC RNA-seq data were available at month 0 and/or at month 3.

### RTS,S/AS01 vaccination is associated with month 3 downregulation of B cell- and monocyte-related BTMs, along with upregulation of T cell-related BTMs

The transcriptional response to RTS,S/AS01 vaccination was assessed in control-stimulated PBMC as well as Ag-stimulated PBMC. Through this approach, we hypothesized that we would see recall responses of Ag-specific T cells activated *in vitro*, as well as responses of other cell types to the secreted cytokines/chemokines. Of note, the sampling schedule at MAL067 was designed for evaluation of acquired immune responses to the vaccine and not ex vivo responses. Our motivation was that in healthy, malaria-naïve adults, the transcriptional response to RTS,S/AS01 has been shown to largely wane by Week 3 post-final dose (16), implying that the majority of the RTS,S/AS01-induced transcriptional changes in this study likely preceded the month 3 sample collection. Three antigens were chosen for stimulation: CSP (peptides covering the CSP region of RTS,S that encodes B-cell and T-cell epitopes), HBS (peptides covering the HBS, also included in the RTS,S vaccine), and AMA1 (a highly immunogenic antigen expressed briefly on hepatocyte-invading *P. falciparum* sporozoites and predominantly on red blood cell-invading *P. falciparum* merozoites, not present in the RTS,S vaccine; included to analyze naturally acquired immunity responses).

Two comparisons were done to characterize the transcriptional response to RTS,S/AS01 vaccination: Comparison (1): comparing gene expression in month 3 samples from RTS,S/AS01 vs comparator recipients (month 3 RTS,S/AS01 vs comparator); and Comparison (2): comparing gene expression in month 3 vs month 0 from RTS,S/AS01 recipients (RTS,S/AS01 month 3 vs month 0). Each comparison has its own advantages: Comparison (1) allows the identification of RTS,S/AS01-specific responses while taking into account other environmental factors to which the children are exposed, such as malaria exposure (albeit malaria transmission intensity was low during the study at both sites). Moreover, the very young ages of the trial participants mean that RTS,S/AS01-induced changes may be confounded with normal developmental changes in participant immune systems, further underscoring the value of Comparison (1), as it does not involve comparison across two different time points. On the other side, an advantage of Comparison (2) is that it takes into consideration each participant’s intrinsic baseline gene expression. Comparison (1) uses data from both infants and children, whereas Comparison (2) can only yield insight into RTS,S/AS01 responses in children (as baseline samples were not collected from infants).

A BTM-based approach was taken to reduce dimensionality, avoid paying a high penalty for multiple testing, and aid results interpretability. For Comparison (1), there were 59 significantly differentially expressed (FDR cutoff ≤ 0.2) BTMs across all antigen stimulation conditions (Figure 2, Table S3a). The vast majority (55) of these BTMs were in vehicle-treated PBMCs, with few differences observed in antigen-stimulated PBMCs (0 significantly differently expressed BTMs in CSP-stimulated PBMC and 8 significantly differentially expressed BTMs in AMA1-stimulated PBMC). The categories with the most differentially expressed BTMs were B cells and T cells: In vehicle-stimulated PBMCs from infants/children who received RTS,S/AS01, 11 T cell-related BTMs were upregulated (vs comparator). We also observed that natural killer (NK) cell and mitochondria-related BTMs were upregulated in vehicle-stimulated PBMC, whereas B cell-related BTMs were downregulated. Counter to our initial expectations, no significant correlations were identified in the CSP-stimulated cells. This result may potentially be explained by the low frequency of CSP-specific T-cells in RTS,S/AS01 vaccinees (e.g. on average, <0.10% of all CD4+ T cells (28)). In AMA1-stimulated cells, surprisingly, the few correlate BTMs that were identified tended to correlate in opposite directions with risk than in the vehicle-stimulated PBMC (i.e. a positive correlate in vehicle-stimulated PBMC yet an inverse correlate in AMA1-stimulated PBMC, and vice versa). We hypothesize that cytokines/chemokines released from activated T cells and their effects on other PBMC may underlie this difference. Alternatively, AMA1 may be eliciting an innate response, which is supported by the findings of Bueno et al. in the context of *P. vivax* (36).

**Figure 2:**
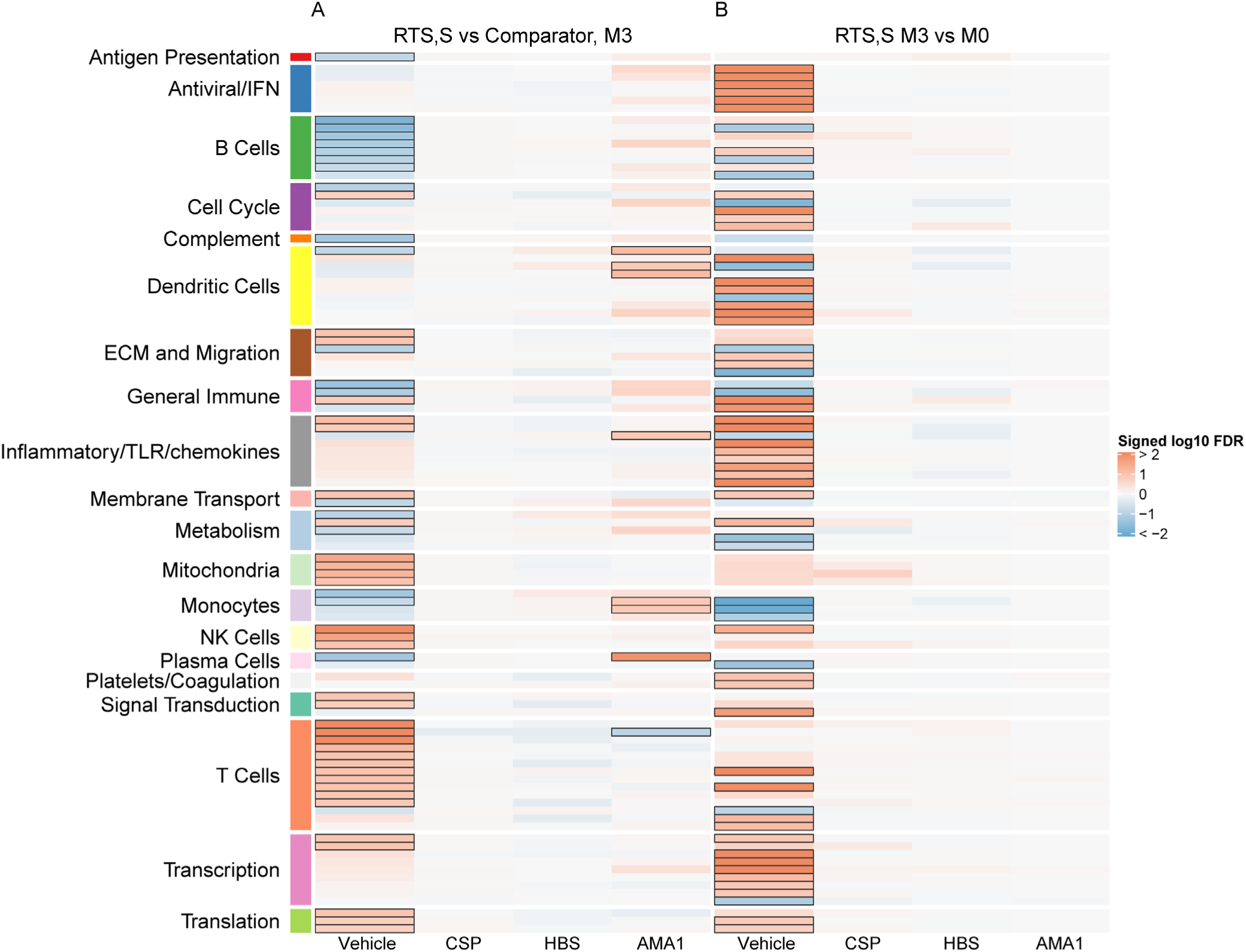
Transcriptional responses and antigen-specific transcriptional responses at one month post-final dose associated with RTS,S/AS01 vaccination. **A)** Comparison 1: month 3 (M3) PBMC, RTS,S/AS01 vs comparator; **B)** Comparison 2: M3 PBMC vs. month 0 (M0) PBMC, RTS,S/AS01 recipients only. Cell color intensity represents the strength of the difference in the relevant comparison, expressed as signed log_10_ false discovery rate (FDR); blood transcriptional modules (BTMs) with significantly different expression (false discovery rate ≤ 0.2) between the two compared groups are outlined in black. Red, higher expression in RTS,S/AS01 recipients vs comparator recipients at M3 (Comparison 1) or higher expression in RTS,S/AS01 recipients at M3 vs M0 (Comparison 2); blue, lower expression in RTS,S/AS01 recipients vs comparator recipients at M3 (Comparison 1) or lower expression in RTS,S/AS01 recipients at M3 vs M0 (Comparison 2). High-level BTM annotation groups are shown in the left-most color bar.

For Comparison (2), only RTS,S/AS01 recipients for whom month 0 and month 3 samples were available were included in the analysis (i.e., children only). There were 67 significantly differentially expressed (FDR cutoff ≤ 0.2) BTMs, all in vehicle-stimulated PBMC (Figure 2, Table S3b). Antiviral/interferon (IFN)-, inflammatory/Toll-like receptor (TLR)/chemokine-, dendritic cell-, and transcription-related BTMs were among those upregulated, whereas B cell- and monocyte-related BTMs were among those downregulated.

The two Comparisons yielded partially divergent results, likely due to the reasons stated above. However, downregulation of monocyte-related BTMs in vehicle-treated PBMC was consistently observed across the two Comparisons: “Enriched in monocytes (II) (M11.0)” was downregulated in both Comparisons, “Enriched in myeloid cells and monocytes (M81)” was downregulated in Comparison (1), and “Monocyte surface signature (S4)” and “Enriched in monocytes (surface) (M118.1)” were downregulated in Comparison (2). Interestingly, monocyte-related BTMs (including these four BTMs) were upregulated at multiple time-points in response to RTS,S/AS01 administration in (16), pointing to differences between the two study populations.

### Monocyte-related RTS,S/AS01 signature BTMs associate with clinical malaria risk

To preserve statistical power in the immune correlates analysis, only BTMs differentially expressed after RTS,S/AS01 vaccination according to Comparison 1 (any stimulation) were down selected. We define these 59 BTMs as the “RTS,S/AS01 signature BTMs”. We next investigated if any of the RTS,S/AS01 signature BTMs were associated with clinical malaria risk in RTS,S/AS01 recipients, by comparing expression of the signature BTMs in cases vs. controls, within each stimulation condition.

In vehicle-stimulated PBMC, five BTMs were significantly differently expressed in RTS,S/AS01 cases vs controls (Figure 3, Table S4): “Myeloid cell enriched receptors and transporters (M4.3)” (up), “Respiratory electron transport chain (mitochondrion) (M219)” (down), “Respiratory electron transport chain (mitochondrion) (M238)” (down), “Enriched in myeloid cells and monocytes (M81)” (up), and “Enriched in monocytes (II) (M11.0)” (up). The results suggest a direct correlation of monocyte activity and an inverse correlation of mitochondrial oxidative phosphorylation 1-month post-primary vaccination with clinical malaria risk. The association of monocyte-related BTMs with risk is consistent with studies reporting a positive correlation between monocyte/lymphocyte ratio and clinical malaria risk and/or severity (37, 38).

**Figure 3:**
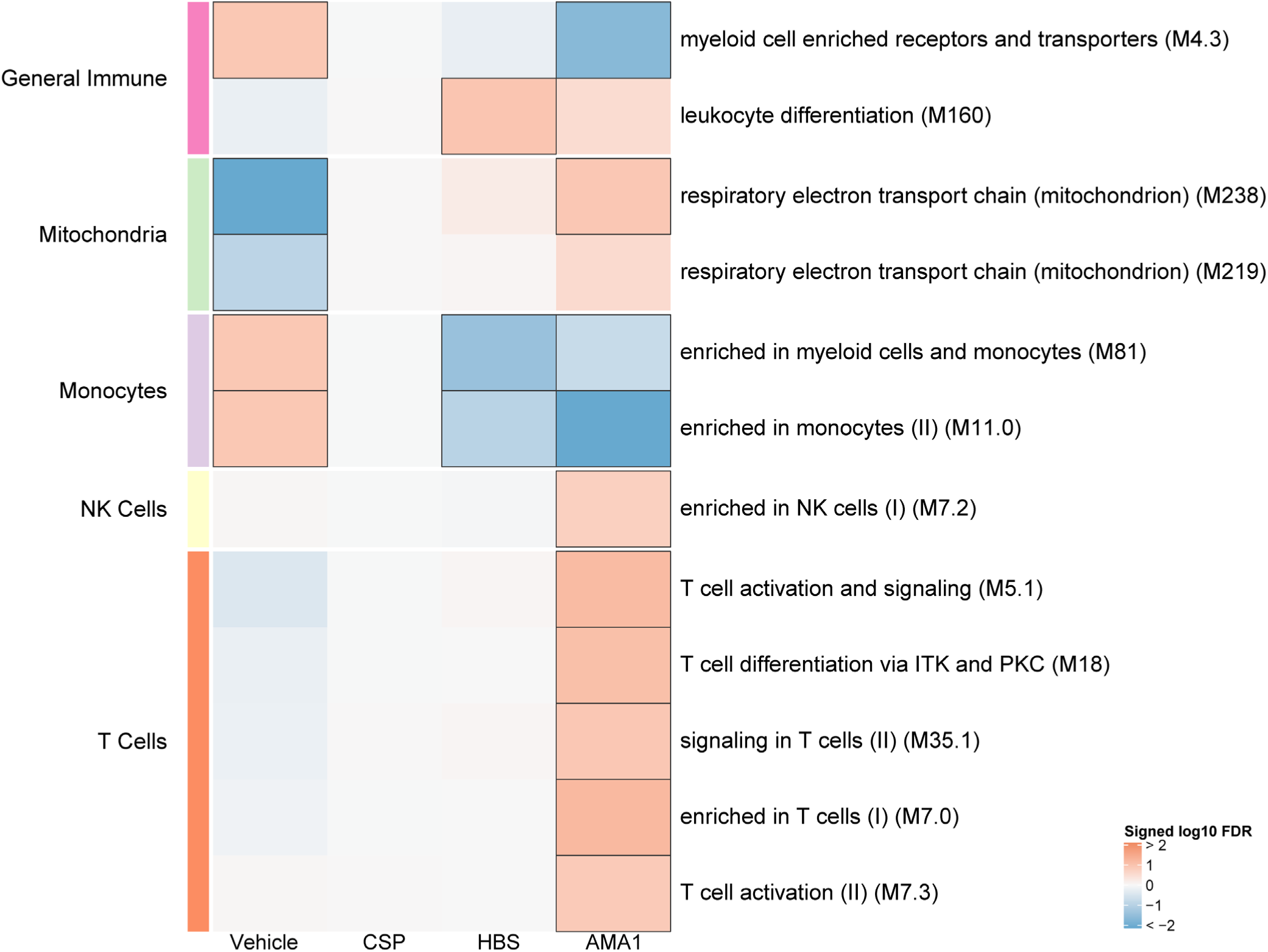
Associations of month 3 levels of RTS,S/AS01 signature blood transcriptional modules (BTMs) with malaria case status in RTS,S/AS01 recipients. Heatmap showing down-selected signature BTMs (Comparison 1) with significantly different expression [false discovery rate (FDR) ≤ 0.2] in month 3 PBMC from RTS,S/AS01 malaria cases vs. non-malaria controls, in at least one stimulation condition. Cell color intensity represents the strength of the difference in the relevant comparison, expressed as signed log_10_ FDR; BTMs with significantly different expression in the comparison are outlined in black. Red, higher expression in RTS,S/AS01 cases vs. controls; blue, lower expression in RTS,S/AS01 cases vs. controls. High-level BTM annotation groups are shown in the left-most color bar.

Antigen-stimulated PBMC also showed transcriptional modules associated with clinical malaria risk. However, these results differed largely from those seen in vehicle-stimulated PBMC. The two monocyte-related BTMs (M81, M11.0) that were directly associated with risk in vehicle-stimulated PBMC were inverse correlates in both HBS- and AMA1-stimulated PBMC. Moreover, five T-cell related BTMs were positive correlates of risk in AMA1-stimulated PBMC (Figure 3).

The same analysis was performed on comparator recipients (Figure S1). For all five BTMs whose levels in vehicle-stimulated PBMC associated either directly or inversely with risk in RTS,S/AS01 recipients (Figure 3), significant correlations were observed in the opposite direction in comparator recipients (Figure S1), suggesting that positive correlations of the three monocyte-related BTMs with risk and inverse correlations of the mitochondria-related BTMs with risk are specific to RTS,S/AS01 recipients.

### Polyfunctional CSP-specific CD4^+^ T-cell responses

In addition to transcriptional changes, our group has shown previously that RTS,S/AS01 vaccination elicits vaccine-specific antibody and cellular responses in African infants and children (e.g. (21, 23, 28)). The polyfunctionality score is a summary measure that encapsulates a participant’s entire Ag-specific T-cell response after vaccination (39). Using data from a pilot study of 179 children (none of whom was a malaria case) at the Manhiça and Bagamoyo sites, Moncunill et al. previously showed that MAL067 RTS,S/AS01 recipients have higher month 3 CSP-specific and HBS-specific CD4+ T-cell polyfunctionality scores than comparator recipients (28). Consistent with this finding, we report that average CSP-specific CD4^+^ T-cell polyfunctionality and magnitude (frequency of CD4^+^ T-cell expressing IL-2 or TNF or CD154) are both higher at month 3 vs. baseline in RTS,S/AS01 vaccine recipients (Figure 4). The few high responders at baseline can likely be attributed to prior malaria exposure. However, there was no difference in average month 3 CSP-specific T-cell response polyfunctionality or magnitude between RTS,S/AS01 cases vs controls (Figure S2).

**Figure 4:**
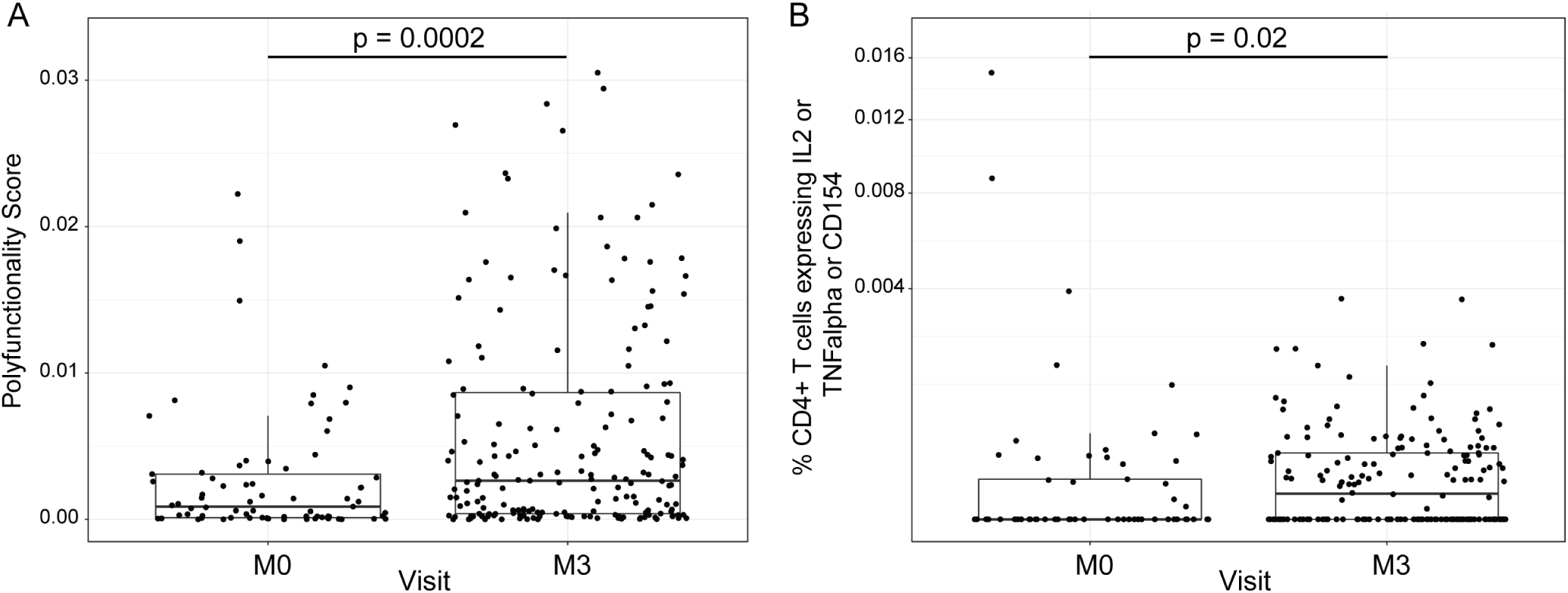
RTS,S/AS01 vaccination elicits polyfunctional T-cell responses. Boxplots show **A)** polyfunctionality score and **B)** magnitude (% CD4+ T cells expressing IL2 or TNF-α or CD154) of CSP-specific CD4+ T-cell responses in RTS,S/AS01 recipients as assessed by intracellular cytokine staining of PBMC collected at month 0 (M0) or at month 3 (M3). Each dot represents a single participant. Data plotted include all available month 0 and month 3 samples, i.e. paired month 0-month 3 samples were not required for plotting. P values represent two-sample test values.

### Month 3 levels of RTS,S/AS01 signature BTMs tend to correlate directly with month 3 IgM antibody responses and inversely with month 3 IgG responses

We next investigated whether month 3 levels of the RTS,S/AS01 signature BTMs were associated with month 3 humoral immune responses in RTS,S/AS01 vaccinees. In vehicle-treated PBMC, both positive and negative associations were seen for multiple antibody variables across functional categories (Figure 5, Table S5). The three antibody variables whose month 3 levels had the strongest positive correlations with transcriptional data were “IgM, LSA1”, “IgM, MSP1 b12 Mad20”, and “IgM, MSP6”, which tended to be positively correlated with month 3 levels of DC-, inflammatory/TLR/chemokine-, and monocyte-related BTMs. In contrast, month 3 levels of “IgG, AMA1 3D7” and “IgG, AMA1 FVO” tended to be inversely correlated with month 3 levels of DC- and monocyte-related BTMs. None of these associations were seen in comparator recipients (Figure S3), suggesting specificity to RTS,S/AS01 receipt, although we note that sample size is smaller which would have reduced statistical power to detect differences. Month 3 levels of cellular variables assessed by polychromatic flow cytometry did not correlate significantly with the month 3 level of any BTM.

**Figure 5:**
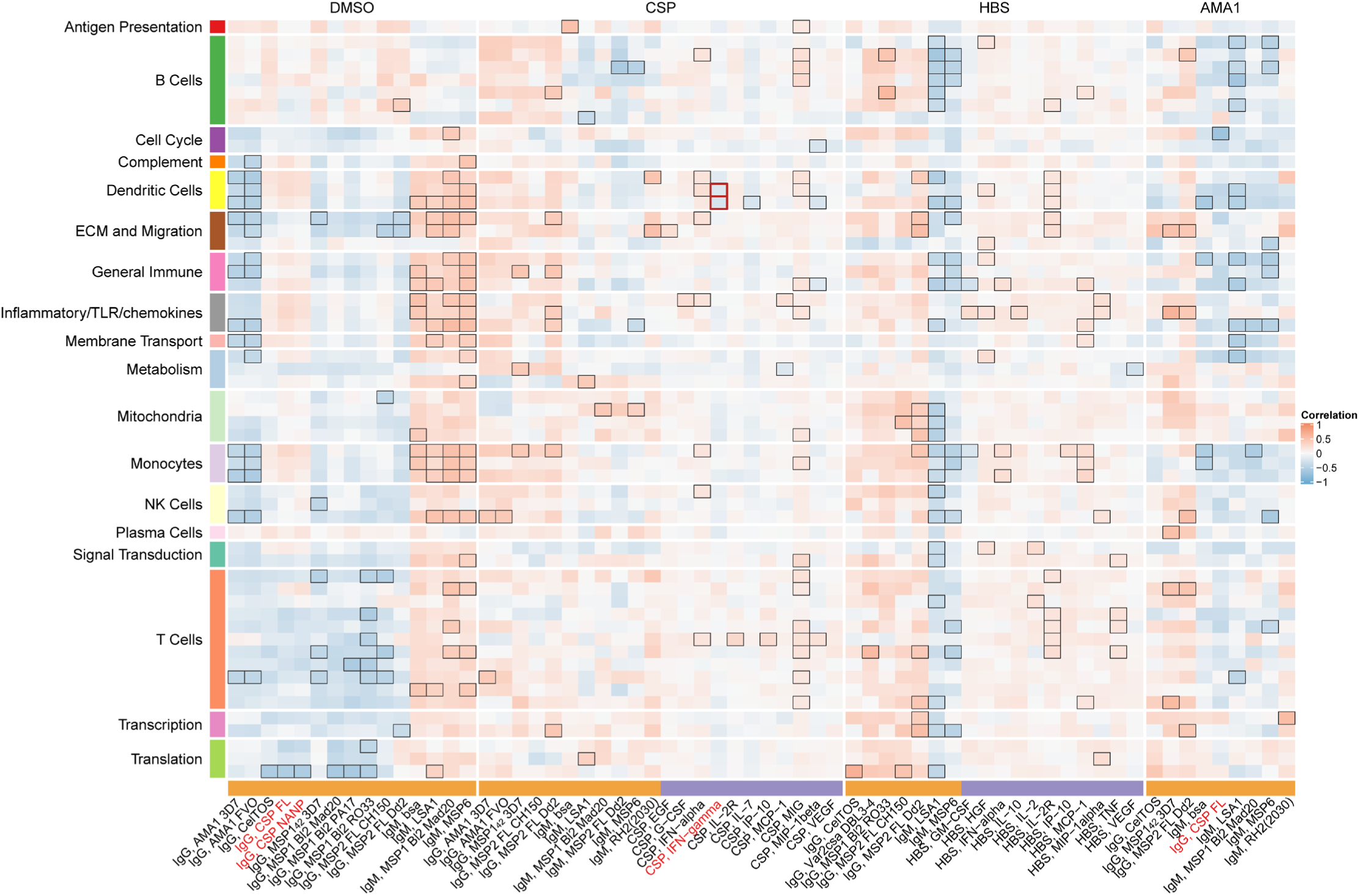
Correlations of month 3 transcriptional and adaptive responses in RTS,S/AS01 vaccine recipients. Heatmap showing correlations between month 3 levels of RTS,S/AS01 signature BTMs and month 3 antibody and cellular responses. Cell color intensity represents the strength of the correlation; BTM/response pairs with significant correlations [false discovery rate (FDR) ≤ 0.2] are outlined in black. Cell color represents correlation direction: red, positive correlation; blue, negative correlation. High-level BTM annotation groups are shown in the left-most color bar. The bottom bar denotes the type of adaptive response analyzed (orange, antibody; purple, cellular). Red font: CSP-specific IFN**-γ** responses have been associated with protection against parasitemia in controlled human malaria infection studies (40, 41); corresponding heatmap squares representing significant BTM/response correlations are bordered in red.

In Ag-stimulated PBMCs, significant correlations of month 3 levels of both antibody and cellular variables with month 3 BTM levels were also observed (Figure 5, Table S5); as above, the correlations were widely spread across BTM functional groupings. For HBS- and AMA1-stimulated PBMCs, antibody variables with the most correlations included “IgM, LSA1” and “IgM, MSP6”. Correlations of month 3 cellular variables with month 3 BTM levels in CSP-stimulated and in HBS-stimulated PBMC were generally positive. Of interest were inverse correlations of month 3 levels of “Resting dendritic cell surface signature (S10)” and “DC surface signature (S5)” with CSP-specific IFN-γ responses (which have been associated with protection in CHMI studies (40, 41) and in the MAL067 study (24)). In general, much fewer associations were seen for comparator recipients (Figure S3). Specifically, the inverse associations of CSP-specific IFN-γ responses with month 3 levels of DC-related BTMs were not seen, suggesting that these associations are RTS,S/AS01-specific.

### Cross-study immune correlates analysis reveals a mostly consistent association in RTS,S/AS01-vaccinees between baseline expression of DC- and monocyte-related BTMs and risk

An important question is whether the results of our analysis of the MAL067 trial, which was conducted in African infants and children in malaria-endemic areas, are generally translatable to other study populations. PBMC transcriptomic data are available for at least three different controlled human malaria infection (CHMI) studies conducted in a quite different study population, i.e. healthy, malaria-naïve adults in the United States. WRAIR 1032 (<u>NCT00075049</u>) randomly assigned participants to receive RTS,S/AS02A or RTS,S/AS01B at months 0, 1, and 2 (40); MAL068 (NCT01366534) randomly assigned participants to receive Ad35.CS.01 at month 0 followed by RTS,S/AS01B at months 1 and 2 (heterologous prime-boost) or RTS,S/AS01B at months 0, 1, and 2 (14); and MAL071 (<u>NCT01857869</u>) randomly assigned participants to receive a full dose of RTS,S/AS01B at months 0, 1, and 2 or a full dose of RTS,S/AS01B at months 0 and 1, followed by a fractional dose at month 7 (42). Importantly, all these trials share a common vaccine arm: one full dose of RTS,S/AS01B at months 0, 1, and 2 (referred to as the “RRR” arm). Microarray data from WRAIR 1032 were analyzed by Vahey et al. (43), microarray data from MAL068 were analyzed by Kazmin et al. (16), and RNA-seq data from MAL068 and MAL071 were analyzed by Du et al. (17).

We performed a cross-study immune correlates analysis where we examined whether the BTMs associated with clinical malaria risk in MAL067 showed similar associations with challenge outcome in each of the three CHMI studies described above. Due to differences in sampling schedules, and the presence of the CHMI challenge (which would complicate results interpretation), we could not compare the exact same month 3 timepoint across studies. We chose instead to compare 21 days post-third dose in MAL068 and in MAL071, i.e. of day of challenge, and 14 days post-third dose in WRAIR 1032, i.e. just before or on day of challenge. We refer to these slightly different post-vaccination time points as “month 3” for simplicity. The month 3 cross-study correlates analysis included BTMs whose month 3 levels (in vehicle-stimulated PBMC) associated with clinical malaria risk in MAL067 RTS,S/AS01B recipients (Figure 3, Table S4) and is shown in Figure 6A. No BTM was consistently associated with malaria risk (or non-protection) across all four studies. The most consistent result was for the monocyte-related BTMs, where month 3 expression of “enriched in monocytes (II) (M11.0)” was significantly associated with risk in 3 of the 4 studies (all except WRAIR 1032), and “enriched in myeloid cells and monocytes (M81)” was significantly associated with risk in MAL067 and MAL071 RRR (Figure 6A, Table S6A).

**Figure 6.**
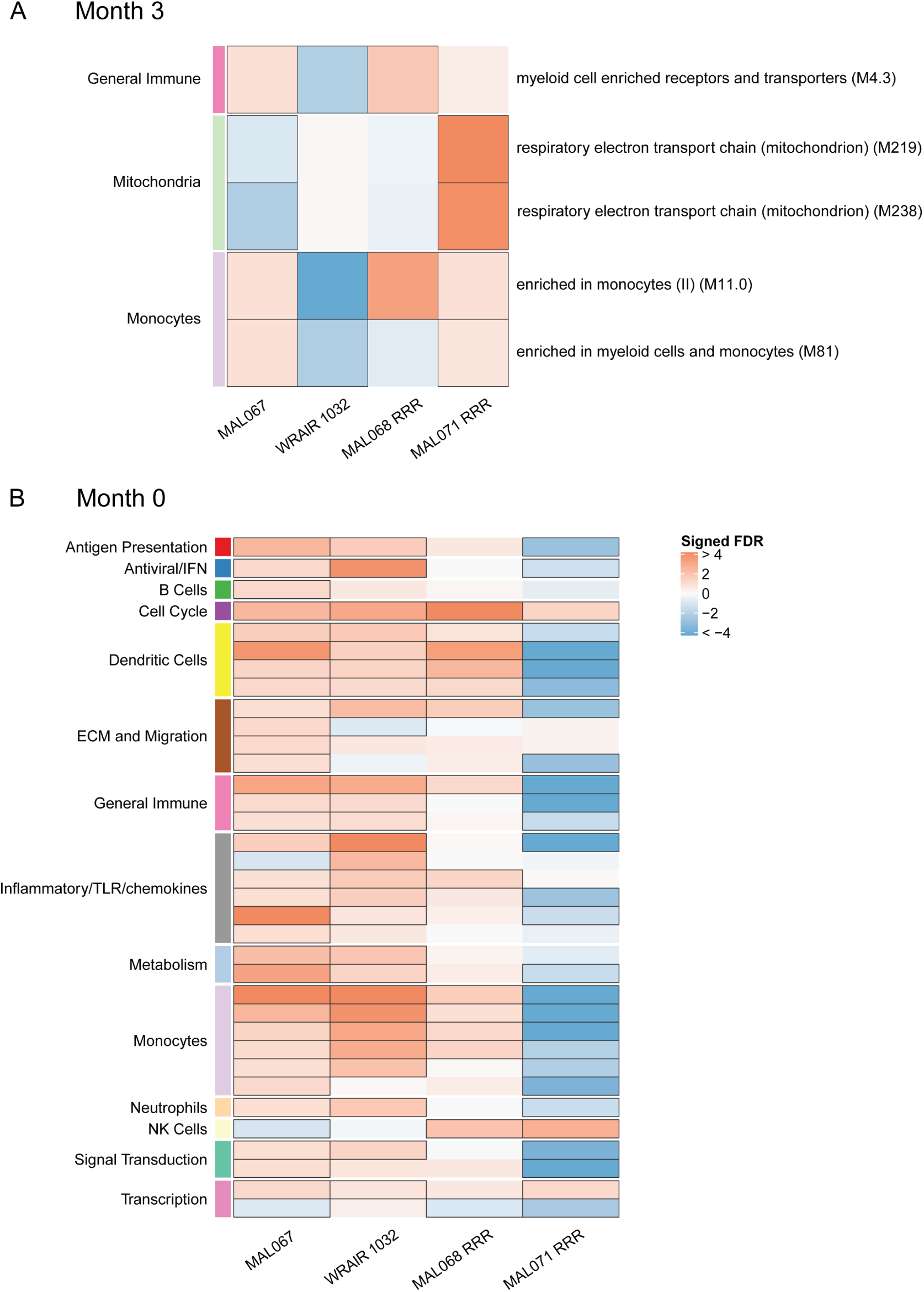
Associations of (A) month 3 or (B) month 0 levels of blood transcriptional modules (BTMs) with malaria case status in MAL067 RTS,S/AS01 vaccine recipients across studies sharing a common month 0, month 1, month 2 RTS,S/AS01 arm. **A)** Heatmap showing the difference in month 3 PBMC BTM expression between RTS,S/AS01 malaria cases vs. non-malaria controls in each of three CHMI studies of the 5 BTMs whose month 3 levels in DMSO-stimulated PBMC associated with malaria case status in MAL067 (Figure 3). Note that “month 3” = 21 days post-final dose in MAL068 and MAL071, and 14 days post-final dose in WRAIR 1032; we refer to these slightly disparate post-vaccination time points as “month 3” for simplicity. BTMs with significantly different expression [false discovery rate (FDR) ≤ 0.2, with adjustment done across the 5 BTMs] are outlined in black. **B)** Heatmap showing the 35 BTMs whose month 0 levels showed significantly different expression in MAL067 RTS,S/AS01 malaria cases vs. non-malaria controls. These 35 BTMs were also examined as potential correlates of challenge outcome in each of the three CHMI studies. Significantly different expression is defined as FDR ≤ 0.2, with adjustment across the 35 BTMs. All data shown are from participants who received the same vaccine regimen: a dose of RTS,S/AS01 at months 0, 1, and 2. MAL067, RNA-seq data from the MAL067 study in African infants and children; WRAIR 1032, Vahey et al. microarray data (43) from the WRAIR 1032 study in malaria-naïve adults (40); MAL068 RRR, RNA-seq data from the RRR arm in the MAL068 study in malaria-naïve adults (14); MAL071 RRR, RNA-seq data from the RRR arm in the MAL071 study in malaria-naïve adults (42). Cell color intensity represents the strength of the difference in the case vs. control comparison, expressed as signed log_10_ false discovery rate (FDR); BTMs with significantly different expression (false discovery rate ≤ 0.2) between the two compared groups are outlined in black. Red, higher expression in RTS,S/AS01 cases vs. controls; blue, lower expression in RTS,S/AS01 cases vs. controls. High-level BTM annotation groups are shown in the left-most color bar.

We next performed the baseline correlates analysis of MAL067 (left-most column, Figure 6B). Compared to the results from the month 3 analysis, the baseline correlates analysis of MAL067 revealed a larger number of BTMs whose month 0 levels (in vehicle-stimulated PBMC) associated (nearly all positively) with clinical malaria risk (Figure 6B, Table S6B). These BTMs spanned many functional categories and were related to DCs, inflammation, and monocytes, among other areas. By far the strongest correlation with risk was with “enriched in monocytes (II) (M11.0)” (FDR = 4.74E-9), followed by “inflammatory response (M33)” (FDR = 1.48E-5) and “resting dendritic cell surface signature (S10)” (FDR = 2.69E-4). Comparing across studies, the greatest similarity seemed to be between MAL067 and WRAIR 1032, which shared 26 common BTMs associated with risk; of these, 12 were also associated with risk in MAL068 RRR. Of note, the two functional annotations whose BTMs most consistently associated with risk across these three studies were “Dendritic cells” (“resting dendritic cell surface signature (S10)”, “complement and other receptors in DCs (M40)”, “DC surface signature (S5)”, and “enriched in dendritic cells (M168)” correlated with risk in all three studies) and “Monocytes” (“enriched in monocytes (I)”, “enriched in monocytes (II)”, “enriched in monocytes “IV”, and “monocyte surface signature (S4)” correlated with risk in all three studies).

In contrast to the multiple positive baseline transcriptional associations with risk in MAL067, WRAIR 1032, and MAL068 RRR, nearly all the month 0 BTMs that associated with challenge outcome in MAL071 RRR were inversely associated with risk, including all the DC- and monocyte-related BTMs seen in the MAL067 signature. We did not find any obvious differences in study population or in sample collection/processing that could explain the differing results. It is also unknown why we see greater consistency in baseline transcriptional associations with risk across studies vs. month 3 transcriptional associations with risk.

Baseline transcriptional associations with month 3 adaptive responses are presented in Figure S4 and Table S7. No significant associations were seen with any antibody responses. Among other functional BTM categories, baseline levels of three monocyte-related BTMs (“Monocyte surface signature (S4)”, “enriched in monocytes (II) (M11.0)”, and “enriched in myeloid cells and monocytes (M81)”), as well as two dendritic-cell-related BTMs (“resting dendritic cell surface signature (S10)” and “DC surface signature (S5)”), all of which associated with risk in the above analysis, showed significant positive correlations with CSP-specific CD4+ T-cell polyfunctionality score.

### Monocyte frequencies tend to be higher at baseline and at month 3 in RTS,S/AS01 vaccinees that develop clinical malaria

The finding that monocyte-related BTMs were expressed significantly higher in RTS,S/AS01 cases vs controls at month 3 in three of the four studies examined (Figure 6A) and at month 0 in three of the four studies examined (Figure 6B) suggested that monocyte frequencies may be higher in cases vs controls at these two timepoints. To determine if the monocyte-related transcriptional differences in PBMC were reflected in monocyte populations, the frequencies of monocytes in PBMC cases and controls were compared using immunophenotyping and flow cytometry. Figure S5A shows that, when automated gating (44) is performed, monocyte frequencies at month 0 and, to a lesser extent at month 3, tend to be higher in cases than in controls for RTS,S/AS01 and comparator recipients combined. However, these differences were not significant (month 0 p=0.131, month 3 p=0.207). When using manual gating, the inflammatory monocyte frequency and inflammatory monocyte/lymphocyte ratio (Figure S5B, S5C) also appeared to be higher in cases vs controls, at both month 3 and baseline, but again these differences were not significant. Thus, these findings do not support our hypothesis that the monocyte-related transcriptional differences in case vs. control PBMC would be reflected in these monocyte populations in cases vs. controls. A potential explanation for why we identified a baseline monocyte transcriptional signature of risk yet did not see an association of baseline monocyte frequency, inflammatory frequency, or inflammatory monocyte/lymphocyte ratio with risk, is that the baseline monocyte transcriptional signature of risk reflects expression changes in the existing circulating monocyte population, rather than an expansion in the circulating monocyte population.

## Discussion

Our main finding is the identification of a baseline blood transcriptional module (BTM) signature, primarily related to dendritic cells, inflammation, and monocytes, that associates with clinical malaria risk in RTS,S/AS01-vaccinated African children. In a cross-study comparison, much of this baseline risk signature – specifically, dendritic cell- and monocyte-related BTMs – was also recapitulated in two of the three CHMI studies in healthy, malaria-naïve adults. Our finding fits into a growing body of evidence that baseline immune status can influence vaccine responses (45). Fourati et al. showed that higher baseline inflammation (as assessed by transcriptomic profiling and flow cytometric analysis of immune cell subset frequencies) was associated with poor antibody response to the hepatitis B vaccine (46). Tsang et al. showed that baseline interferon signaling was robustly correlated with maximum fold change (post-influenza vaccination to baseline) in influenza-specific antibody titer (47). The HIPC Consortium identified a baseline inflammatory gene signature that was associated with higher antibody responses to influenza vaccine in younger participants, yet lower antibody responses in older participants (48). Kotliarov et al. identified baseline transcriptional signatures that predicted antibody responses to the live attenuated yellow fever vaccine and to the trivalent inactivated influenza vaccine (49). Moreover, Hill et al. reported that increased baseline frequencies of plasmablasts and of circulating T follicular helper cells were associated with higher post-RTS,S/AS01 vaccination antibody titers (50). The mechanisms underlying the association found in the present study of baseline monocyte and inflammation-related gene expression with malaria risk in RTS,S/AS01 recipients are unknown, but this association does have potentially important implications, if this association is validated in additional studies of African children in malaria-endemic areas (as our cross-study comparison only looked at CHMI studies in malaria-naïve adults). For instance, an intriguing possibility is that prevaccination administration of anti-inflammatory drugs before vaccine inoculation may potentially reduce baseline inflammation and hence improve RTS,S/AS01-mediated protection. This strategy has been successfully followed in improving antibody responses to the 2012 seasonal influenza vaccine (Agrippal inactivated influenza vaccine) in the elderly (51), albeit it required a 6-week treatment course of the mTOR inhibitor RAD001. While such a treatment course would pose significant logistical challenges in our context, the general approach could potentially become more feasible if new (or existing) anti-inflammatory drugs are found that would require shorter administration periods; there is currently substantial interest towards taking such an approach to improve vaccine response e.g. in older adults (52).

The role of inflammatory monocytes in vaccine-immunity is an area of active research, however, the association of monocyte activity with clinical malaria risk is consistent with studies that have reported a positive correlation between monocyte to lymphocyte (ML) ratio and clinical malaria risk and/or severity (37, 38). Of note, the association between clinical malaria risk and ML ratio was found to be independent of both age and antibodies to parasite blood-stage antigens (38). Warimwe et al. reported that the ML ratio modifies the VE of RTS,S, where VE is lower at higher ML ratios (53). The results of Warimwe et al. raised the hypothesis that inflammatory monocytes may inhibit RTS,S-induced protective adaptive responses. Though the setting is different, the findings that monocytes inhibit T cell proliferation (54, 55), T cell activation (55), and B cell responses (56) in various mouse models of viral infection provide a potential basis for this hypothesis. Further studies are needed to determine whether such a inhibitory mechanism is at play in RTS,S vaccine immunity and whether strategies for modulating monocyte populations via chemokine receptor antagonists, as proposed by (55), could help boost RTS,S efficacy. However, here we found that baseline expression of monocyte-related BTMs was positively associated with post-vaccination CSP-specific polyfunctional CD4+ T cell responses. A potential explanation for this association may be more efficient antigen presentation to T cells upon vaccination in individuals with higher baseline expression of these monocyte-related BTMs. The association of month 3 levels of monocyte-related BTMs with risk may be independent from polyfunctional T cells, which we did not find to associate with risk or protection in this work.

Our finding of the association of monocyte signatures with malaria risk seem to contradict the results of a recent study in which we found that monocyte BTMs were associated with protection in RTS,S/AS01 vaccinated children and infants (57). However, our previous monocyte-protection association was observed after analyzing gene expression levels in CSP-stimulated, background-corrected (i.e. after subtracting expression in vehicle-stimulated) PBMC (57), whereas the monocyte-risk association described in this study was observed after analyzing gene expression levels in vehicle-stimulated PBMC. Here, we did not detect BTMs associated with the response to RTS,S/AS01 vaccination or with protection when analyzing CSP-stimulated PBMC. This result is not surprising, given the low frequency of CSP-specific T-cells in RTS,S/AS01 vaccinees (e.g. on average, <0.10% of all CD4+ T cells (28)) and the number of T-cell non-responders based on the ICS data reported in this manuscript. In fact, in the previous study we could identify BTMs associated with RTS,S/AS01 vaccination in CSP-stimulated PBMC, but no differentially expressed genes were detected (57) and we had used different stimulation conditions and microarrays instead of RNAseq.

It is perhaps counterintuitive – considering that the RTS,S/AS01 vaccine does not contain AMA1 – that we observed a small number of BTMs associated with the response to RTS,S/AS01 vaccination and with clinical malaria risk when analyzing AMA1-stimulated PBMC. To explain this result, we refer the reader to our previous work that showed that RTS,S/AS01 vaccination alters antibody responses to antigens not contained in the RTS,S/AS01 vaccine (22). RTS,S/AS01 recipients received partial protection from the RTS,S/AS01 vaccine, leading possibly to decreased *P. falciparum* parasite load and/or exposure (infection). We hypothesize that the AMA1 stimulation activated T cells that had been previously primed by prior exposure to *P. falciparum* and that RTS,S/AS01 recipients had fewer primed T cells due to decreased *P. falciparum* infection (via partial RTS,S/AS01 protection), providing a potential explanation for the transcriptional differences in AMA1-stimulated PBMC between RTS,S/AS01 vs comparator recipients.

We next discuss some limitations of our study. First, in malaria-naïve adults, the transcriptional response to the third RTS,S/AS01 dose has been shown to peak at Day 1 post-injection, with some decline by Day 6 and approximately 90% of the response having waned by Day 21 (16). Therefore, it is likely that the sampling scheme in this study (one month post-final dose) misses the majority of the transcriptional response to RTS,S/AS01. Future studies with dense, early post-vaccination PBMC sampling could be useful for further investigating RTS,S transcriptional immune correlates. Second, PBMCs were stimulated on site and then frozen. As each site performed the procedure separately, this renders our data susceptible to batch effects. However, a standardized SOP and shared reagents were used, decreasing the possibility of such effects. Moreover, an advantage of on-site stimulation of fresh PBMC is that it avoids the decrease in cell viability, and potential loss of detection of Ag-specific cells, that may have occurred if PBMC had been frozen, thawed, and then stimulated at a central location. Third, there was confounding between age and location. As all infants were from Manhiça and the majority of children were from Bagamoyo, it was not possible to examine the impact of age or clinical trial site on RTS,S/AS01 transcriptional response. Fourth, as only patrolling cell subsets are present in PBMC, we were unable to detect potential signals from T cells, B cells, NK cells, and macrophages localized to an infection site including skin and liver or the immune memory compartment localized in secondary lymphoid organs.

Despite these limitations, our study also has a number of strengths. For example, while excellent work has already been done to interrogate transcriptional responses to RTS,S/AS01 vaccination in healthy, malaria-naïve adults (including densely sampled early post-vaccination sampling timepoints to capture innate responses) and to identify molecular correlates of RTS,S/AS01-mediated protection against clinical malaria after CHMI in malaria-naïve adults (16, 17, 43, 58), in our study we examined transcriptional responses to RTS,S/AS01 vaccination in infants and children in malaria-endemic areas. This feature is a strength of our study, as 1) infants in particular have relatively immature immune systems (59), making it likely that infants (and younger children) mount different vaccine responses than adults (60); 2) infants and children are especially susceptible to malaria-related morbidity and mortality, making them the target population for this and other malaria vaccines; and 3) continual exposure to *P. falciparum*, as occurs in endemic areas, influences naturally acquired immunity, which in turn interacts with immunity conferred by RTS,S vaccination (22). Related to this, another advantage of our study is the use of a comparator group which allows to discern the effect of the vaccine from environmental exposures including *P. falciparum* and age. As participants in the study are very young, significant development of their immune systems occurs throughout the duration of the study, meaning that such changes could potentially be confounded with vaccine-induced immune changes.

While it will be necessary to perform follow-up studies at more sites and with larger sample sizes to validate the baseline transcriptional signature associated with malaria risk identified here, our study raises interesting hypotheses related to the relationship of inflammation and RTS,S/AS01-mediated protection and suggests potential strategies to explore for augmenting RTS,S/AS01 VE.

## Data Availability

Data are available from the corresponding authors upon reasonable request.

## Acknowledgments

We are very grateful to study participants, their families, and vaccine trial site field and lab staff. We thank the Phase 3 trial sites PIs Salim Abdulla, Pedro Alonso, Jahit Sacarlal, and Pedro Aide; the investigators involved in the generation of immunology data used here, including providers of antigens for antibody assays (Itziar Ubillos, Marta Vidal, Alfons Jimenez, Ruth Aguilar, Diana Barrios, Laura Puyol, Aintzane Ayestaran, Luis Izquierdo, David Cavanagh, James Beeson, David Lanar, Vir Chauhan, Chetan Chitnis, Deepak Gaur, Evelina Angov, Benoit Gamain, Ross Coppel); the MAL067 Vaccine Immunology Consortium investigators and Working Groups; and Fergal Duffy for valuable comments on the manuscript. We thank GlaxoSmithKline Biologicals S.A. for their support in the conduct of the MAL067 study.

## Supplementary Material

### Supplementary Methods

Quality control of RNA sequencing data: Multidimensional scaling (MDS) plots as implemented in the plotMDS function of the limma package were used to visualize variability across samples and identify potential abnormalities (batch effects, outliers) or patterns of biological interest (association within experimental factors).

---

**Figure S1.**
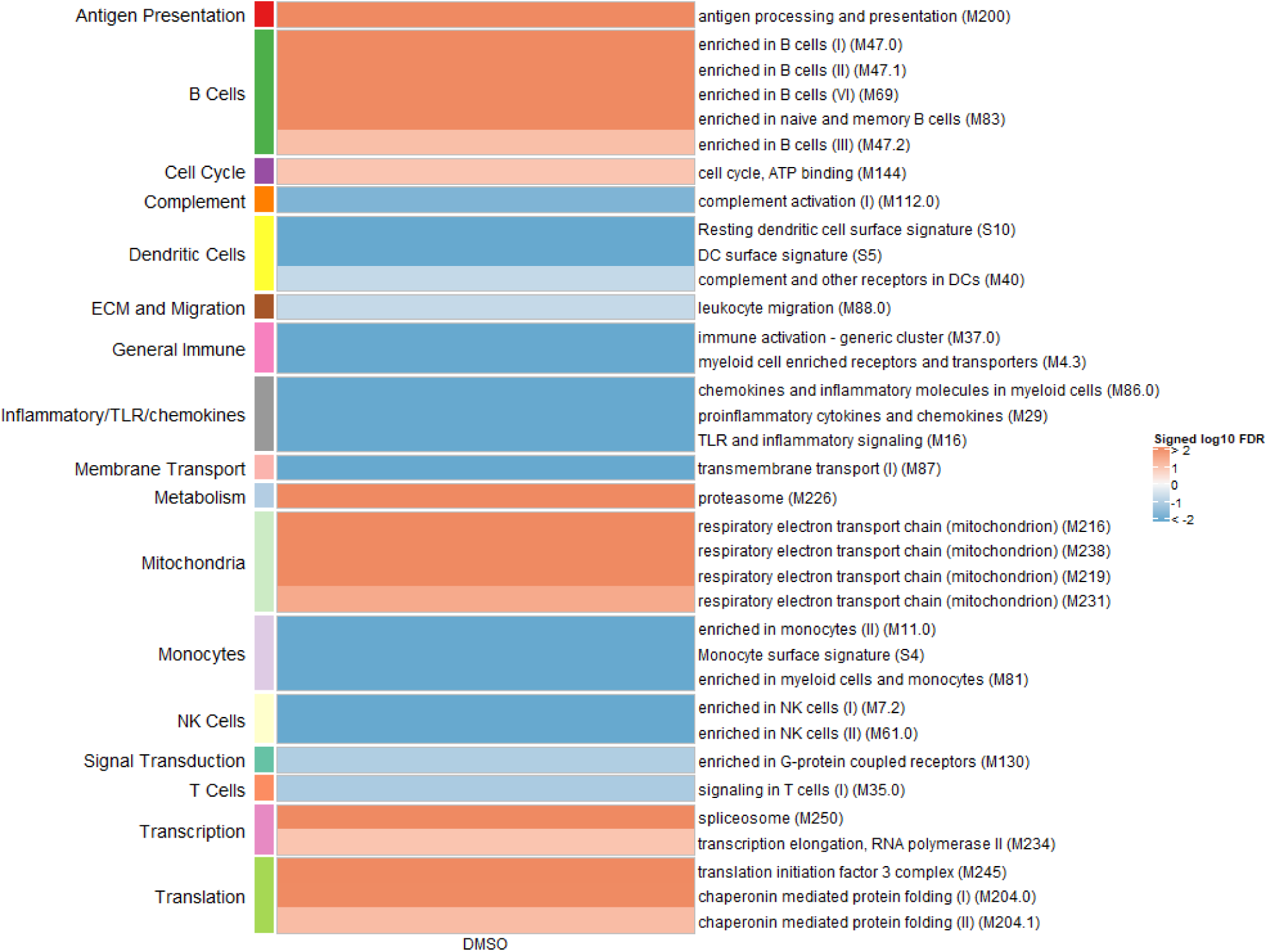
Associations of month 3 levels of RTS,S/AS01 signature blood transcriptional modules (BTMs) with malaria case status in comparator recipients. Heatmap showing which of the 59 down-selected RTS,S/AS01 signature BTMs (Comparison 1) showed significantly different expression [false discovery rate (FDR) ≤ 0.2] in month 3 PBMC between comparator cases vs. non-malaria controls in DMSO-stimulated PBMC. Analyses were also done with Ag-stimulated PBMC but are not shown here as none of the results were significant (only DMSO had any significant BTMs). Cell color intensity represents the strength of the difference in the relevant comparison, expressed as signed log_10_ FDR. Red, higher expression in comparator cases vs. controls; blue, lower expression in comparator cases vs. controls. High-level BTM annotation groups are shown in the left-most color bar.

**Figure S2:**
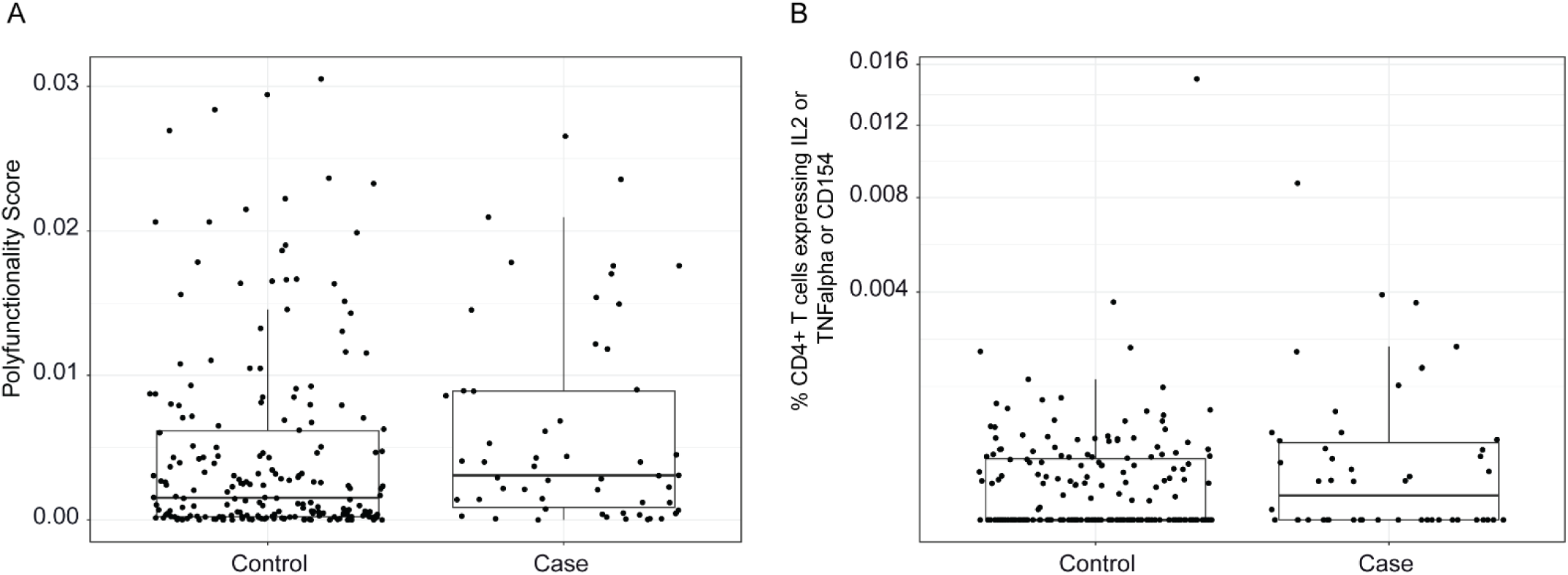
A) Polyfunctionality score and B) magnitude of CSP-specific CD4+ T-cell responses in RTS,S/AS01 vaccine recipients at month 3, stratified by case (clinical malaria)-control status.

**Figure S3:**
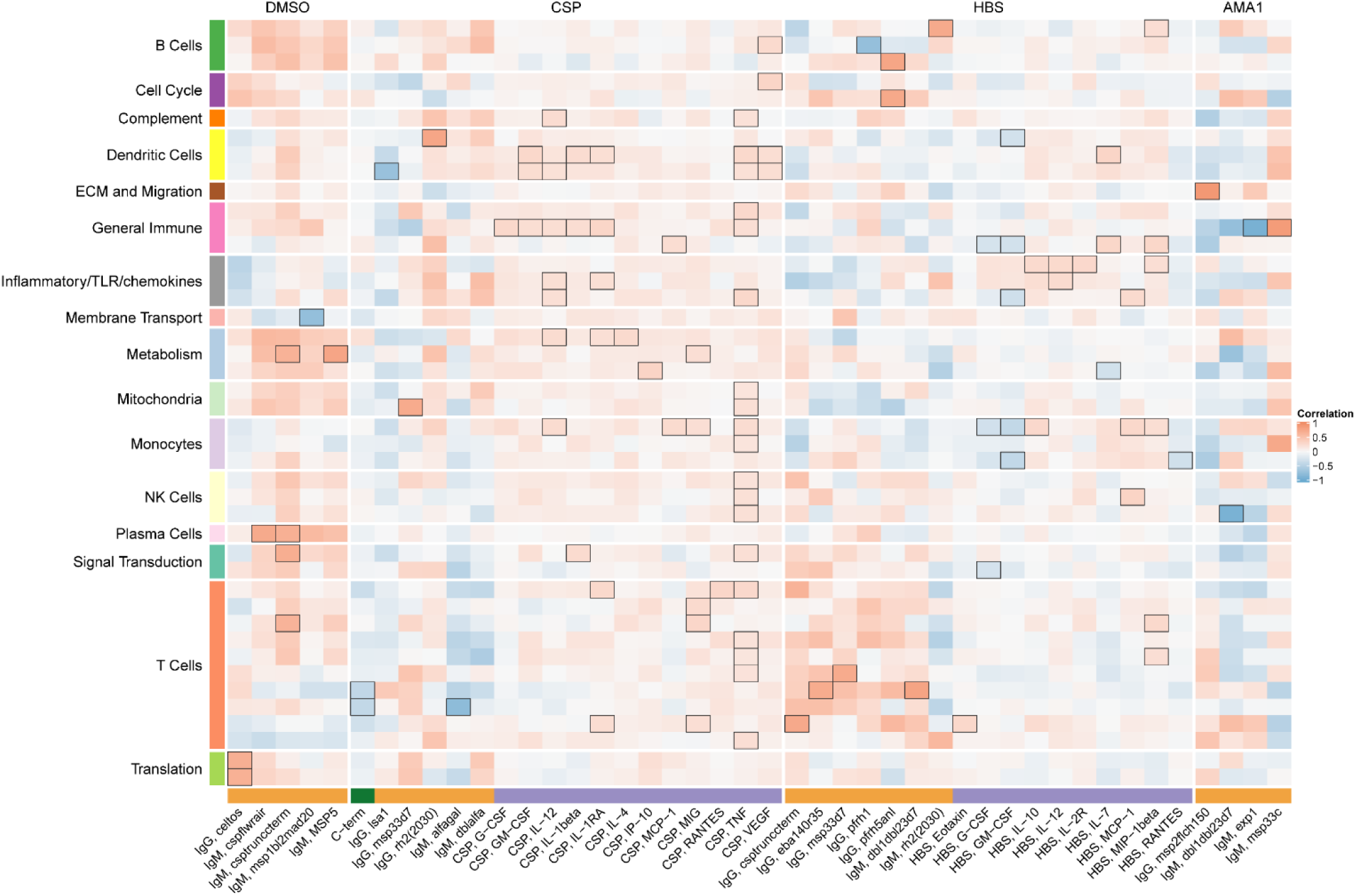
Correlations of transcriptional and adaptive responses in comparator vaccine recipients. Heatmap showing significant correlations between month 3 levels of RTS,S/AS01 signature BTMs and month 3 antibody and cellular responses in comparator recipients. Cell color intensity represents the strength of the correlation; BTM**/**response pairs with significant correlations [false discovery rate (FDR) ≤ 0.2] are outlined in black. Cell color represents correlation direction: red, positive correlation; blue, negative correlation. High-level BTM annotation groups are shown in the left-most color bar. The bottom bar denotes the type of adaptive response analyzed (orange, antibody; purple, cellular). Green bar, IAVI measurements.

**Figure S4.**
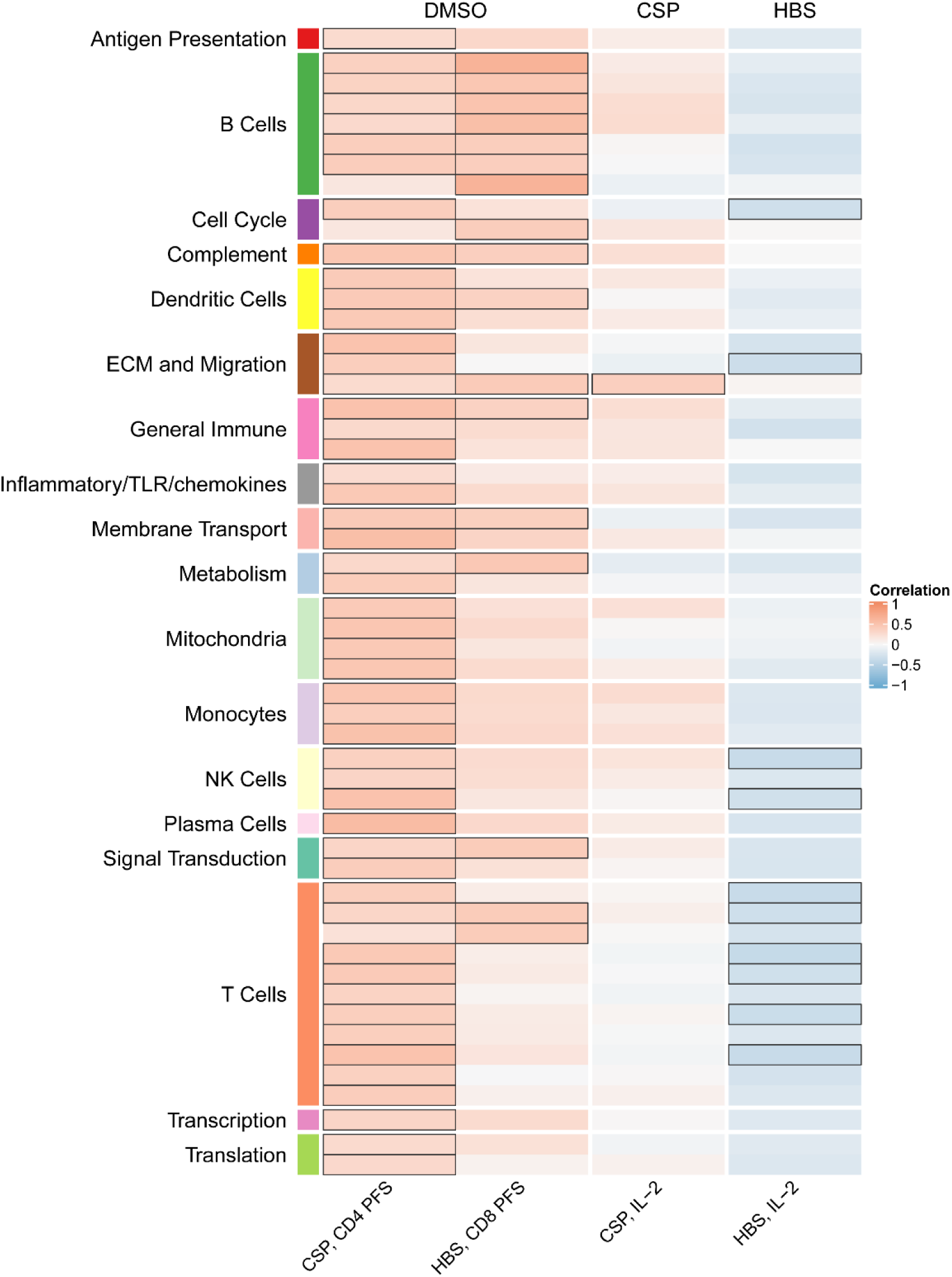
Correlations of Month 0 blood transcriptional module (BTM) expression with month 3 adaptive responses in RTS,S/AS01 vaccine recipients. Heatmap showing significant correlations between month 0 levels of RTS,S/AS01 signature BTMs and month 3 cellular responses (no significant correlations were seen with any month 3 antibody responses). Cell color intensity represents the strength of the correlation; BTM/response pairs with significant correlations [false discovery rate (FDR) ≤ 0.2] are outlined in black. Cell color represents correlation direction: red, positive correlation; blue, negative correlation. High-level BTM annotation groups are shown in the left-most color bar.

**Figure S5.**
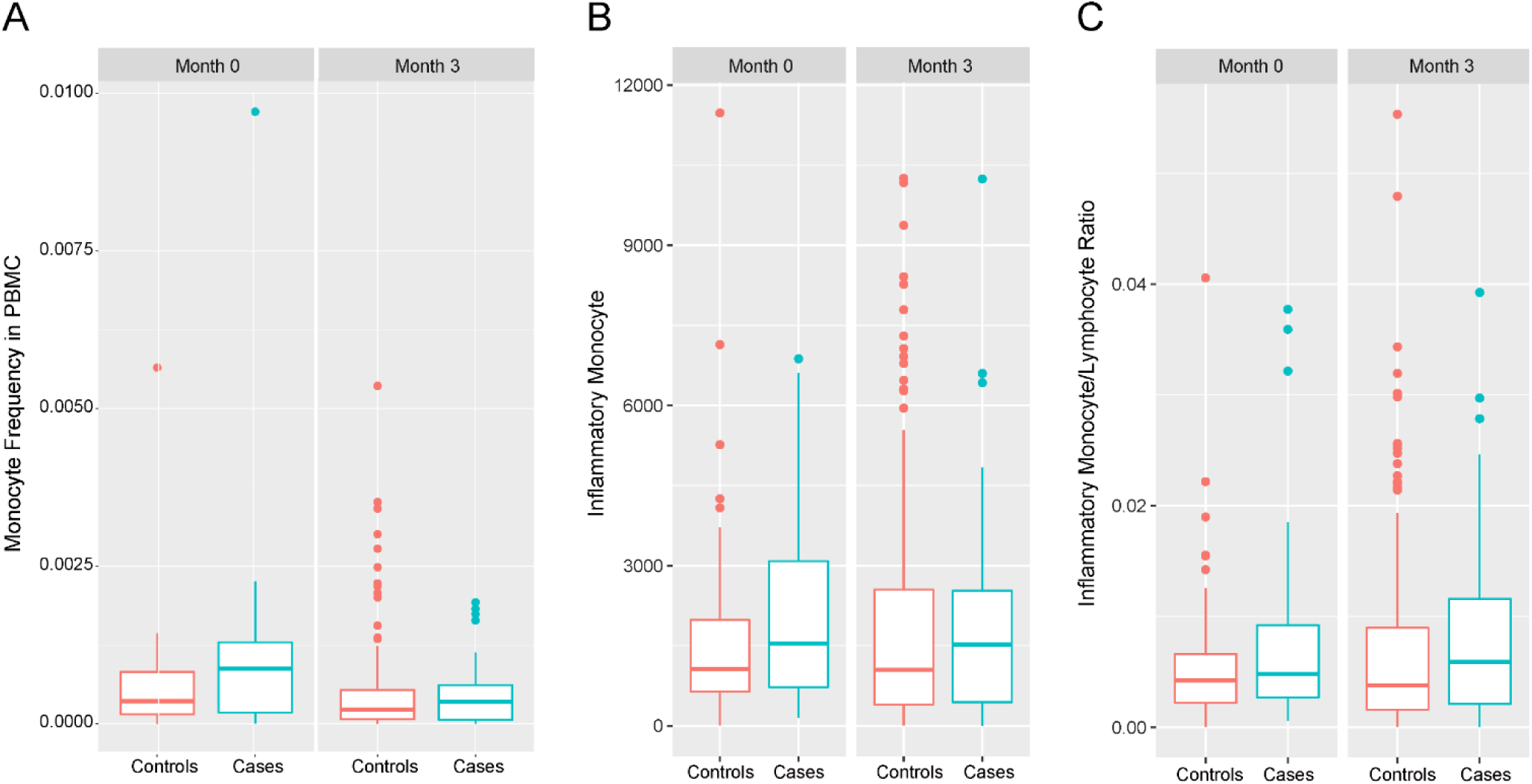
RTS,S/AS01 cases tend to have higher monocyte frequencies than controls, at both baseline and at month 3. (A) Monocyte frequencies in PBMC, (B) inflammatory monocyte counts in PBMC, and (C) ratios of inflammatory monocytes to lymphocytes. Data are shown separately in cases and in controls (RTS,S/AS01 recipients only) in each panel. (A) was assessed using FAUST; (B) and (C) were assessed using De Rosa flow immunophenotyping. “Inflammatory monocytes” refers to “HLA-DR+ CD14+CD16++” cells.

**Table S1.**
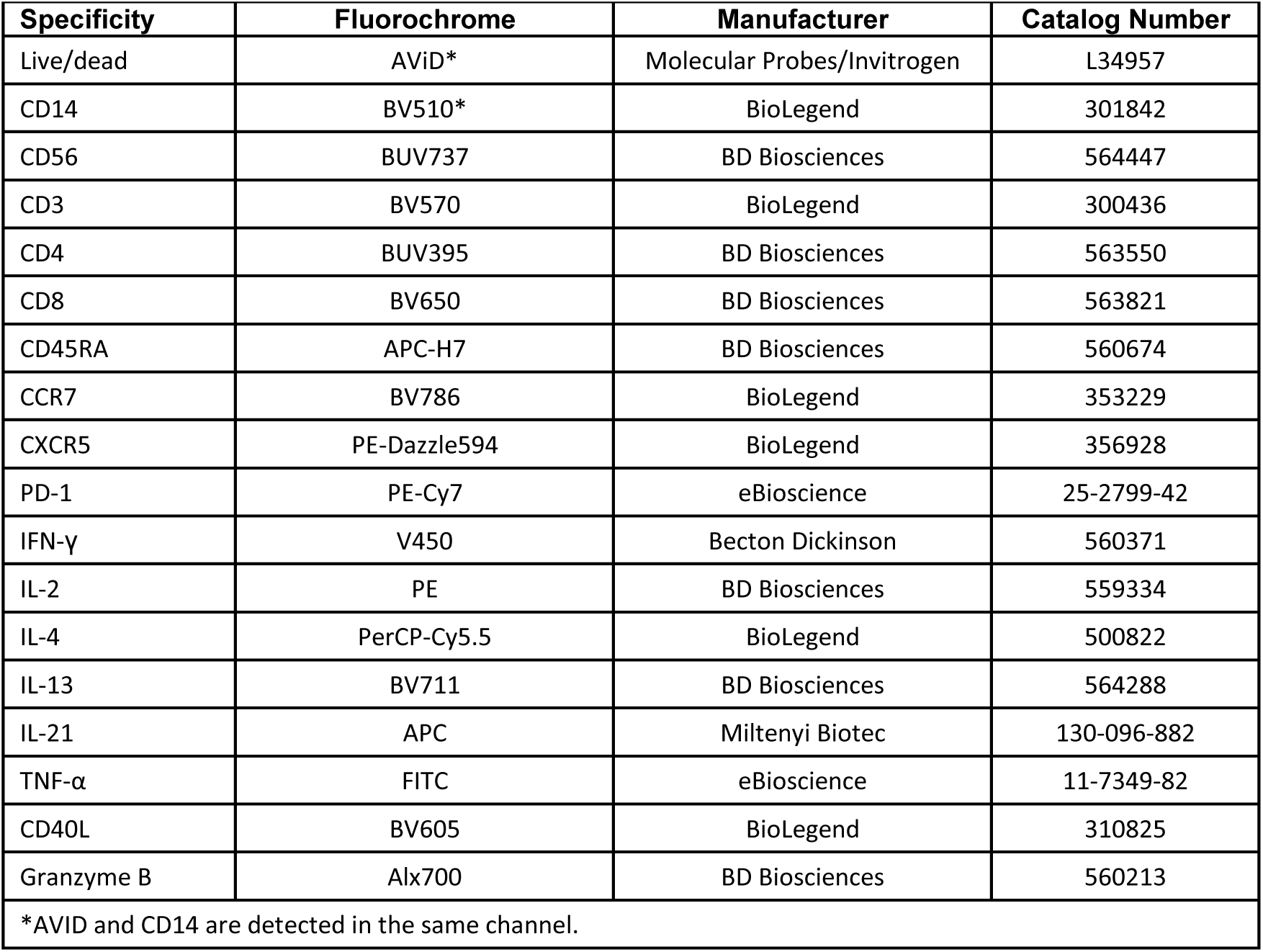
Antibody reagents included in the ICS panel.

**Table S2.**
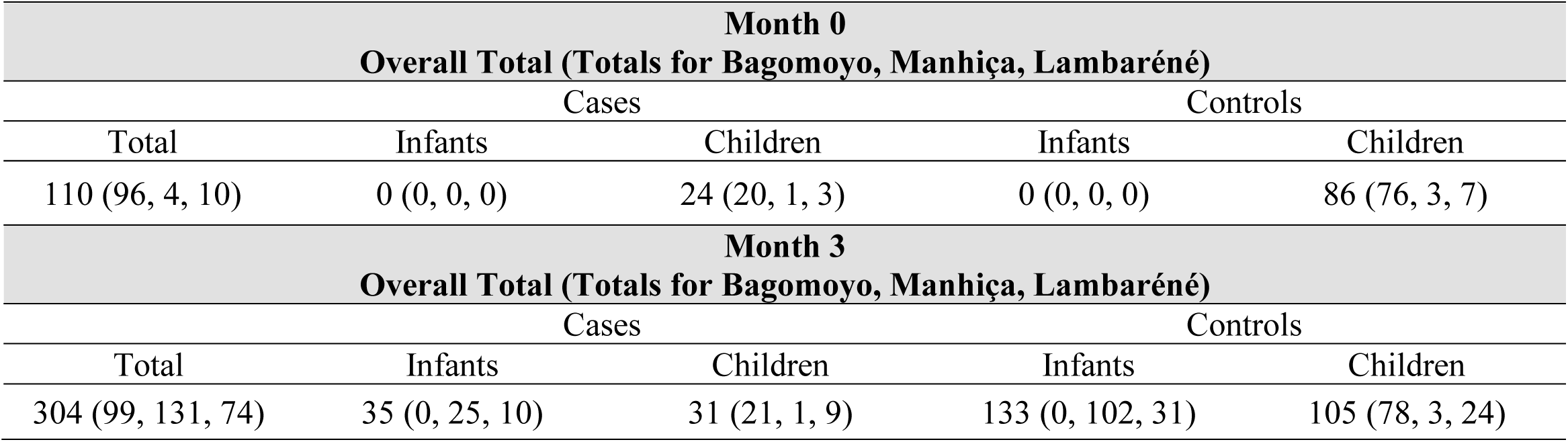
Numbers, age group, and case-control status of RTS,S/AS01 recipients by site for whom month 0 and/or month 3 PBMC samples were included in the ICS/immunophenotyping analysis.

(Tables S3 through S9 are uploaded as separate Excel files)

## Supplementary Table Legends, Tables S3 through S9

Table S3. List of BTMs, p values, and FDRs for Comparison 1) RTS,S/AS01 vs comparator recipients at month 3 and Comparison 2) RTS,S/AS01 recipients at month 3 vs month 0.

Table S4. List of all BTMs tested for significantly different expression in RTS,S/AS01 cases vs controls at month 3, along with stimulation, p value, and FDR results for each Comparison.

Table S5. List of BTMs whose month 3 levels correlated significantly with at least one month 3 adaptive response variable in RTS,S/AS01 vaccinees, along with stimulation, variable details, p value, and FDR results.

Table S6. List of BTMs whose month 3 or month 0 levels had significantly different expression in RTS,S/AS01 cases vs. controls in MAL067, along with p values and FDR results when testing the MAL067 sets of correlate BTMs for significantly different expression in cases vs. controls in the WRAIR 1032, MAL068 RRR, and MAL071 RRR studies.

Table S7. List of BTMs whose month 0 levels correlated significantly with at least one month 3 adaptive response variable, along with stimulation, variable details, p value, and FDR results.

Table S8. List of all BTMs with significantly different expression in comparator cases vs controls at month 3, along with p value and FDR results for each Comparison. Only vehicle had any significant (FDR ≤ 0.2) BTMs.

Table S9. List of BTMs whose month 3 levels correlated significantly with at least one month 3 adaptive response variable in comparator recipients, along with stimulation, variable details, p value, and FDR results.

