## Supplementary Material for "A baseline transcriptional signature associates with clinical malaria risk in RTS,S/AS01-vaccinated African children"

### Supplementary Methods

Quality control of RNA sequencing data:  Multidimensional scaling (MDS) plots as implemented in the plotMDS function of the limma package were used to visualize variability across samples and identify potential abnormalities (batch effects, outliers) or patterns of biological interest (association within experimental factors).

**Figure S1. Associations of month 3 levels of RTS,S/AS01 signature blood transcriptional modules (BTMs) with malaria case status in comparator recipients.** Heatmap showing which of the 59 down-selected RTS,S/AS01 signature BTMs (Comparison 1) showed significantly different expression [false discovery rate (FDR) ≤ 0.2] in month 3 PBMC between comparator cases vs. non-malaria controls in DMSO-stimulated PBMC. Analyses were also done with Ag-stimulated PBMC but are not shown here as none of the results were significant (only DMSO had any significant BTMs). Cell color intensity represents the strength of the difference in the relevant comparison, expressed as signed $\log_{10}$ FDR. Red, higher expression in comparator cases vs. controls; blue, lower expression in comparator cases vs. controls. High-level BTM annotation groups are shown in the left-most color bar.

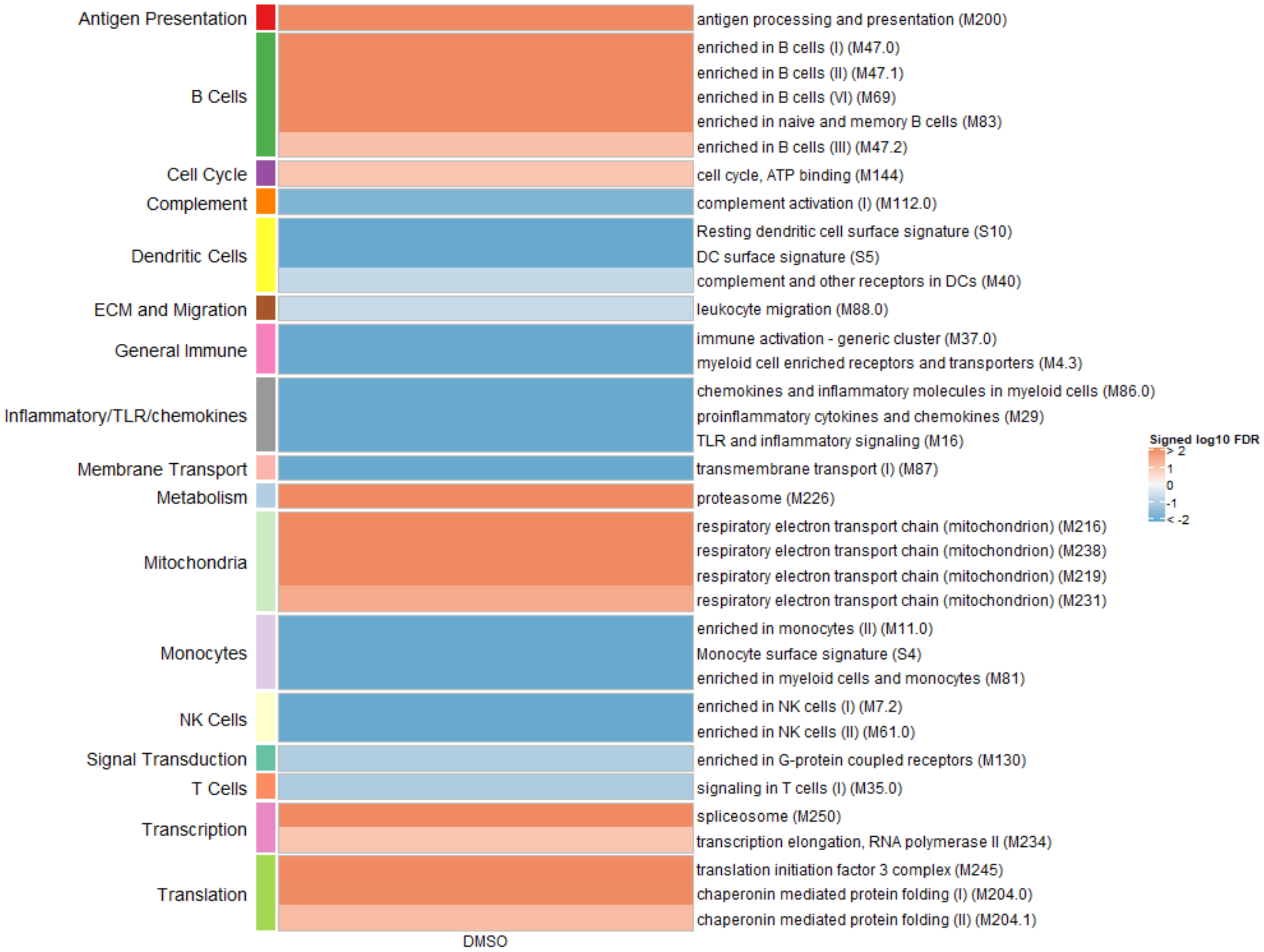

**Figure S2: A) Polyfunctionality score and B) magnitude of CSP-specific C D4+ T-cell responses in RTS,S/AS01 vaccine recipients at month 3, stratified by case (clinical malaria)-control status.**

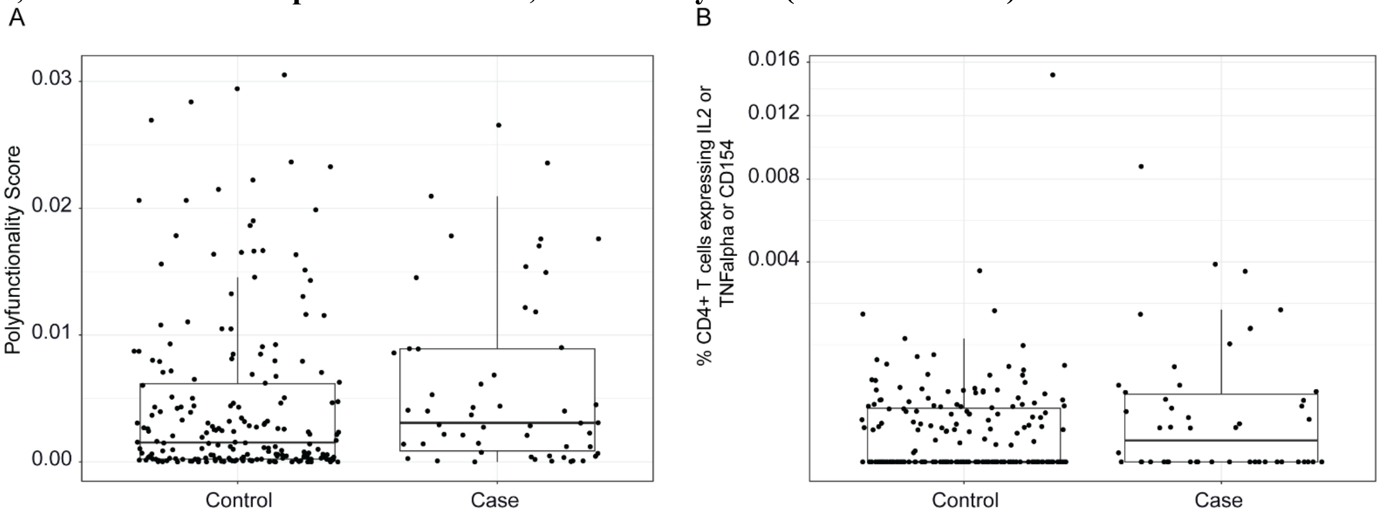

**Figure S3: Correlations of transcriptional and adaptive responses in comparator vaccine recipients.** Heatmap showing significant correlations between month 3 levels of RTS,S/AS01 signature BTMs and month 3 antibody and cellular responses in comparator recipients. Cell color intensity represents the strength of the correlation; BTM/response pairs with significant correlations [false discovery rate (FDR) ≤ 0.2] are outlined in black. Cell color represents correlation direction: red, positive correlation; blue, negative correlation. High-level BTM annotation groups are shown in the left-most color bar. The bottom bar denotes the type of adaptive response analyzed (orange, antibody; purple, cellular). Green bar, IAVI measurements.

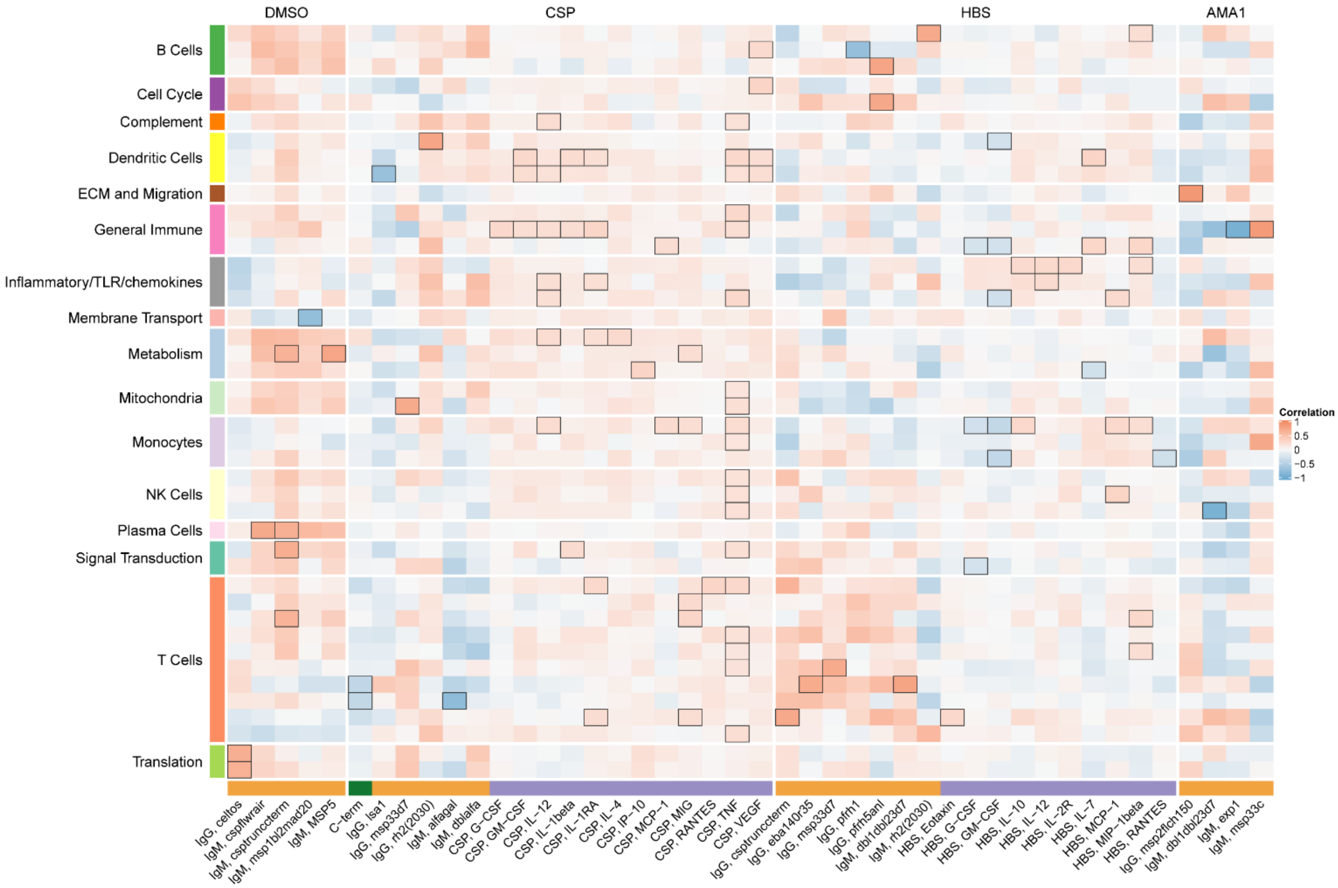

**Figure S4. Correlations of Month 0 blood transcriptional module (BTM) expression with month 3 adaptive responses in RTS,S/AS01 vaccine recipients.** Heatmap showing significant correlations between month 0 levels of RTS,S/AS01 signature BTMs and month 3 cellular responses (no significant correlations were seen with any month 3 antibody responses). Cell color intensity represents the strength of the correlation; BTM/response pairs with significant correlations [false discovery rate (FDR) ≤ 0.2] are outlined in black. Cell color represents correlation direction: red, positive correlation; blue, negative correlation. High-level BTM annotation groups are shown in the left-most color bar.

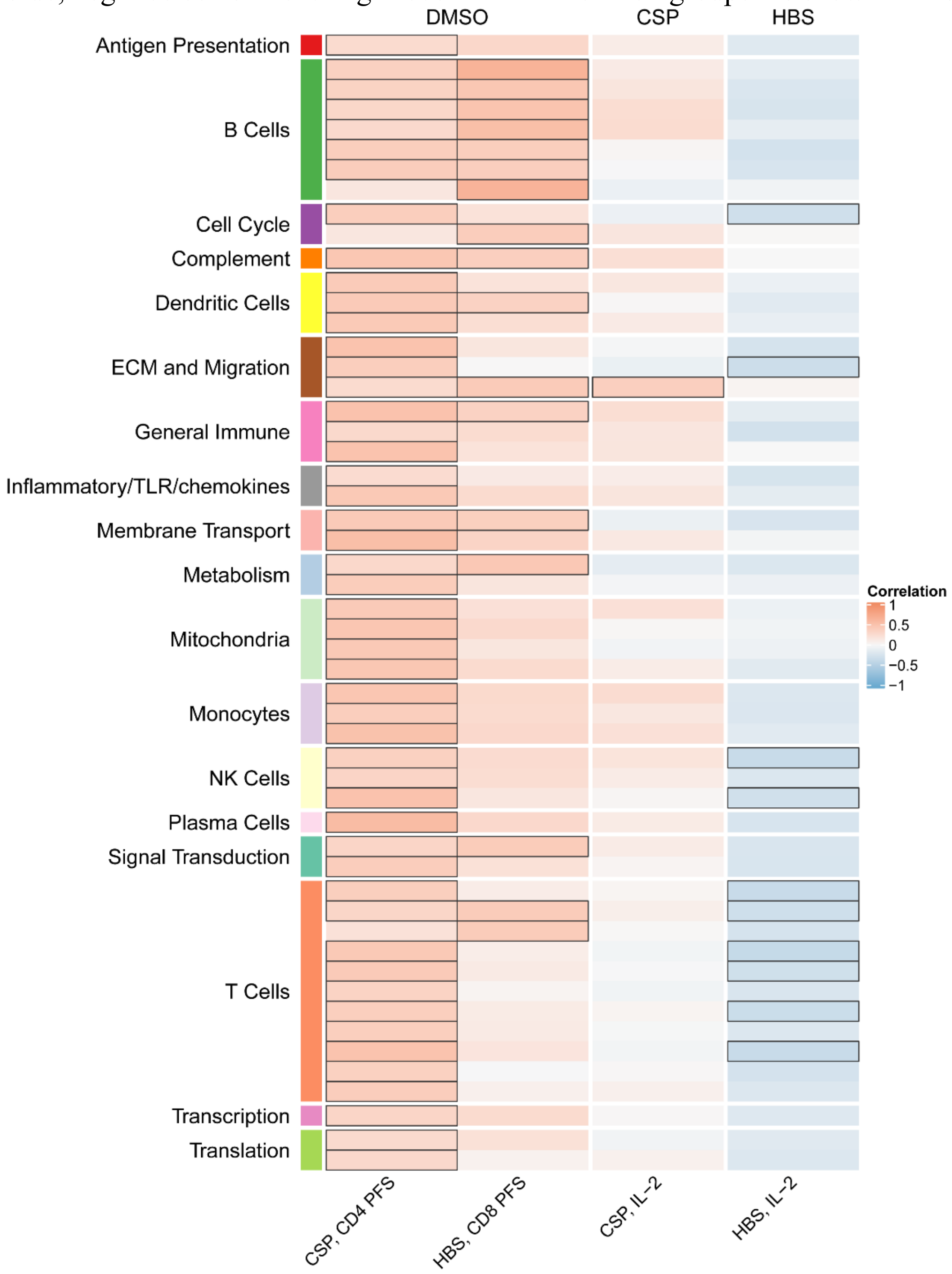

**Figure S5. RTS,S/AS01 cases tend to have higher monocyte frequencies than controls, at both baseline and at month 3.** (A) Monocyte frequencies in PBMC, (B) inflammatory monocyte counts in PBMC, and (C) ratios of inflammatory monocytes to lymphocytes. Data are shown separately in cases and in controls (RTS,S/AS01 recipients only) in each panel. (A) was assessed using FAUST; (B) and (C) were assessed using De Rosa flow immunophenotyping. "Inflammatory monocytes" refers to "HLA-DR+ CD14+CD16++" cells.

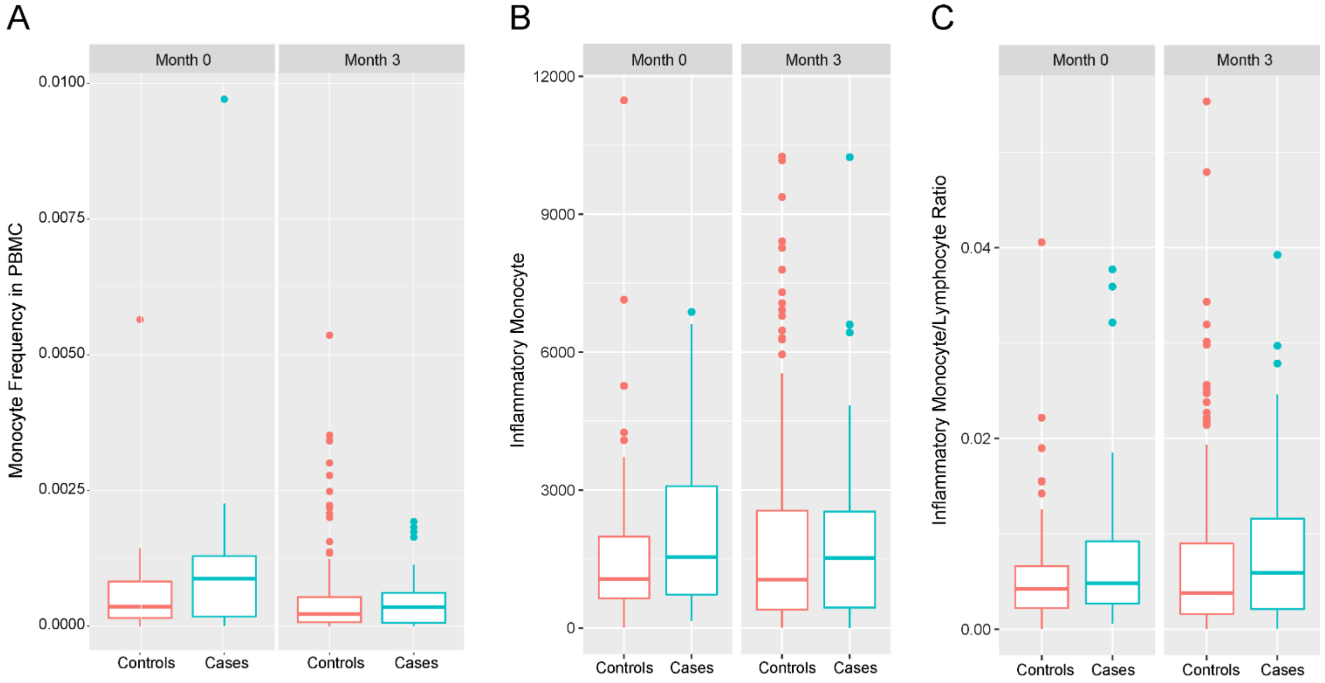

**Table S1. Antibody reagents included in the ICS panel.**

| Specificity | Fluorochrome | Manufacturer | Catalog Number |
|---|---|---|---|
| Live/dead | AViD* | Molecular Probes/Invitrogen | L34957 |
| CD14 | BV510* | BioLegend | 301842 |
| CD56 | BUV737 | BD Biosciences | 564447 |
| CD3 | BV570 | BioLegend | 300436 |
| CD4 | BUV395 | BD Biosciences | 563550 |
| CD8 | BV650 | BD Biosciences | 563821 |
| CD45RA | APC-H7 | BD Biosciences | 560674 |
| CCR7 | BV786 | BioLegend | 353229 |
| CXCR5 | PE-Dazzle594 | BioLegend | 356928 |
| PD-1 | PE-Cy7 | eBioscience | 25-2799-42 |
| IFN-γ | V450 | Becton Dickinson | 560371 |
| IL-2 | PE | BD Biosciences | 559334 |
| IL-4 | PerCP-Cy5.5 | BioLegend | 500822 |
| IL-13 | BV711 | BD Biosciences | 564288 |
| IL-21 | APC | Miltenyi Biotec | 130-096-882 |
| TNF-α | FITC | eBioscience | 11-7349-82 |
| CD40L | BV605 | BioLegend | 310825 |
| Granzyme B | Alx700 | BD Biosciences | 560213 |
| *AVID and CD14 are detected in the same channel. | | | |

**Table S2. Numbers, age group, and case-control status of RTS,S/AS01 recipients by site for whom month 0 and/or month 3 PBMC samples were included in the ICS/immunophenotyping analysis.**

| **Month 0<br>Overall Total (Totals for Bagomoyo, Manhiça, Lambaréné)** | | | | |
|---|---|---|---|---|
| | Cases | | Controls | |
| Total | Infants | Children | Infants | Children |
| 110 (96, 4, 10) | 0 (0, 0, 0) | 24 (20, 1, 3) | 0 (0, 0, 0) | 86 (76, 3, 7) |
| **Month 3<br>Overall Total (Totals for Bagomoyo, Manhiça, Lambaréné)** | | | | |
| | Cases | | Controls | |
| Total | Infants | Children | Infants | Children |
| 304 (99, 131, 74) | 35 (0, 25, 10) | 31 (21, 1, 9) | 133 (0, 102, 31) | 105 (78, 3, 24) |

(Tables S3 through S9 are uploaded as separate Excel files)

**Supplementary Table Legends, Tables S3 through S9**

Table S3. List of BTMs, p values, and FDRs for Comparison 1) RTS,S/AS01 vs comparator recipients at month 3 and Comparison 2) RTS,S/AS01 recipients at month 3 vs month 0.

Table S4. List of all BTMs tested for significantly different expression in RTS,S/AS01 cases vs controls at month 3, along with stimulation, p value, and FDR results for each Comparison.

Table S5. List of BTMs whose month 3 levels correlated significantly with at least one month 3 adaptive response variable in RTS,S/AS01 vaccinees, along with stimulation, variable details, p value, and FDR results.

Table S6. List of BTMs whose month 3 or month 0 levels had significantly different expression in RTS,S/AS01 cases vs. controls in MAL067, along with p values and FDR results when testing the MAL067 sets of correlate BTMs for significantly different expression in cases vs. controls in the WRAIR 1032, MAL068 RRR, and MAL071 RRR studies.

Table S7. List of BTMs whose month 0 levels correlated significantly with at least one month 3 adaptive response variable, along with stimulation, variable details, p value, and FDR results.

Table S8. List of all BTMs with significantly different expression in comparator cases vs controls at month 3, along with p value and FDR results for each Comparison. Only vehicle had any significant ($FDR \leq 0.2$) BTMs.

Table S9. List of BTMs whose month 3 levels correlated significantly with at least one month 3 adaptive response variable in comparator recipients, along with stimulation, variable details, p value, and FDR results.
